# AI-assisted continuous-time modelling of metastatic breast cancer reveals subtype-specific spatiotemporal organ interactions

**DOI:** 10.64898/2026.06.15.26355684

**Authors:** Marc Vaisband, Gabriel Rinnerthaler, Simon Peter Gampenrieder, Nadine Binder, Christian Fridolin Singer, Thamer Sliwa, Florian Roitner, Christopher Hager, Petra Pichler, Simon Udovica, Rupert Bartsch, Sonja Heibl, Daniel Egle, Clemens A. Schmitt, August Felix Zabernigg, Margit Sandholzer, Johannes Andel, Renate Pusch, Richard Greil, Jan Hasenauer

**Affiliations:** Bonn Center for Mathematical Life Sciences; Life & Medical Sciences Institute, University of Bonn, Bonn, Germany; Division of Clinical Oncology, Department of Internal Medicine, Medical University of Graz, Graz, Austria; Department of Internal Medicine III with Haematology, Medical Oncology, Haemostaseology, Infectiology and Rheumatology, Cancer Research Laboratory, University Hospital Salzburg, Paracelsus Medical University Salzburg, Salzburg, Austria; Institute of General Practice/Family Medicine, Faculty of Medicine and Medical Center, University of Freiburg, Freiburg, Germany; Department of Obstetrics and Gynecology and Comprehensive Cancer Center, Medical University of Vienna, Vienna, Austria; Department for Internal Medicine, Haematology and Internal Oncology, LKH Hochsteiermark, Leoben, Austria; Department of Internal Medicine II, Hospital Braunau, Braunau, Austria; Breast Center Dornbirn, Dornbirn, Austria; Department for Internal Medicine 1, University Hospital St. Pölten, St. Pölten, Austria; Department of Medicine I, Clinic Ottakring, Vienna, Austria; Department of Medicine I, Division of Oncology, Medical University of Vienna, Vienna, Austria; Department of Internal Medicine IV, Klinikum Wels-Grieskirchen GmbH, Wels, Austria; Department of Gynaecology, Medical University Innsbruck, Innsbruck, Austria; Department of Hematology and Internal Oncology, Med Campus III, Kepler University Medical Center, Johannes Kepler Universität Linz, Linz, Austria; Department of Internal Medicine, County Hospital Kufstein, Kufstein, Austria; Department of Internal Medicine II, Academic Teaching Hospital Feldkirch, Feldkirch, Austria; Department of Internal Medicine II, Pyhrn-Eisenwurzen Klinikum Steyr, Steyr, Austria; Internal Medicine I for Hematology with Stem Cell Transplantation, Hemostaseology and Medical Oncology, Ordensklinikum Linz Barmherzige Schwestern – Elisabethinen, Linz, Austria; Salzburg Cancer Research Institute-Laboratory of Immunological and Molecular Cancer Research (SCRI-LIMCR) Oncomathematics Group, Salzburg, Austria; Salzburg Cancer Research Institute-Center for Clinical Cancer and Immunology Trials (SCRI-CCCIT), Salzburg, Austria; Austrian Group for Medical Tumor Therapy (AGMT)

**Author notes:** These authors contributed equally (Marc Vaisband, Gabriel Rinnerthaler). These authors jointly supervised this work (Richard Greil, Jan Hasenauer).

## Abstract

Metastatic breast cancer is one of the leading causes of premature mortality among women worldwide. A major barrier to optimal care is the marked heterogeneity in both the temporal dynamics of metastatic spread and the organ-specific spatial distribution of metastases. Existing analyses do not adequately capture this complexity, as they either neglect temporal dependencies or assume independence between metastasic sites. As a result, it remains unclear how established metastases influence subsequent organ-specific dissemination. We address this question using patient-level longitudinal trajectories from a large multicentre real-world metastatic breast cancer registry, combined with an AI-assisted disease-progression modelling framework based on continuous-time Markov chains that represent combinations of metastatic sites and the non-uniform and practice-driven timing of radiologic response assessments, as encountered in routine clinical care. We present a stochastic model determined by progression rates, which are parameterised to capture baseline organ-specific transition risks, patient-level covariates, and pairwise inter-organ interaction effects. High-dimensional treatment information is incorporated using an large language model based encoding. We find that metastatic spread follows non-independent, subtype-specific spatiotemporal patterns, with subtype-specific inter-organ interaction patterns that shape progression. Visceral metastases, particularly lung and liver metastasis, are associated with an increased hazard of subsequent brain metastasis, with effects varying across hormone receptor-positive, HER2-positive, and triple-negative subtypes. Together, these findings define a clinically relevant spatiotemporal architecture of metastatic progression in breast cancer. This framework enables refined mechanism-informed risk stratification and provides a data-driven rationale for targeted and risk-adapted – rather than symptom-triggered – surveillance strategies.

## 1 Introduction

Breast cancer (BC) is the most common cancer in women and remains the second leading cause of cancer-related mortality among the female population worldwide [WMM+17]. While the majority of patients are diagnosed at an early stage, up to one-third develop metastatic disease [MOS+24], which remains the principal driver of mortality. One of the challenges in metastatic breast cancer lies in the marked heterogeneity of both the spatial distribution and temporal evolution of disease spread. A better understanding of the underlying dynamics may help tailor treatment strategies.

Large-scale clinical, pathological, and molecular studies have established that the metastatic pattern of breast cancer is profoundly shaped by tumour-intrinsic characteristics. Foremost among these are the subtypes based on hormone receptor (HR) and HER2 status [BWS+17; KYW+10; RTS+18]. HR-positive (HR+) tumours, particularly luminal subtypes, most frequently metastasize to bone and are often associated with a relatively indolent course [BWS+17; RTS+18; DCB+24]. In contrast, HER2-positive (HER2+) and triple-negative breast cancers (TNBC) display an increased propensity for early and aggressive visceral metastases, especially to the lung and brain. TNBC patients, for instance, are nearly four times as likely to develop lung and brain metastases within five years after diagnosis compared to non-TNBC patients [DHT+09].

Beyond receptor status, histological characteristics play a critical role in shaping disease trajectory. Histological grade acts as a key prognostic factor; high-grade tumours are associated with a higher incidence of visceral spread and poorer outcomes, whereas low-grade tumours more frequently remain confined to bone [GWH+22; RTS+18]. Together, these factors define distinct but highly variable disease trajectories, suggesting that metastatic spread may follow structured, yet patient-specific spatiotemporal trajectories.

These observed associations have informed clinical risk stratification and surveillance strategies. However, their interpretation in real-world data remains challenging. First, longitudinal patient trajectories are typically sparse and unevenly sampled, reflecting the less standardized timing of staging assessments in routine clinical practice, and are further influenced by evolving treatment landscapes, which complicates the separation of intrinsic disease biology from treatment effects. Improved systemic control may prolong survival and thereby increase the observed incidence of late-occurring metastases, (e.g. brain metastases in luminal BC), without necessarily reflecting true organ-specific tropism. Second, analyses are subject to censoring and competing risks, particularly death, which may obscure downstream metastatic events. As a result, apparent differences in organ-specific risk may reflect differences in survival rather than underlying biological propensity. Third, most existing approaches implicitly assume independence between metastatic sites, thereby precluding the identification of inter-organ dependencies, where established metastases may actively modulate subsequent dissemination.

To accurately capture these complexities, modelling methods must account for a broad range of covariates as well as the innate stochasticity of the disease. Markov chains have proven useful in describing probabilistic transitions between metastatic sites. For instance, discrete-time Markov chains have been used to show how the site of initial metastasis shapes subsequent progression [NMV+15] and to predict bone metastases in inflammatory breast cancer [FMC+19]. Complementary prognostic and machine-learning approaches, including HER2DX signatures in early HER2-positive breast cancer and recent deep learning models integrating high-dimensional clinical and molecular data, further highlight the potential of computational methods for patient-specific metastatic risk prediction [DCB+24; XFL+25]. Mechanistic mathematical models have also been developed to capture key steps of the metastatic cascade and the spatial–temporal complexity of tumour–microenvironment interactions [FLB+19]. However, standard discrete-time analyses often fail to account for the non-uniform and practice-driven timing of radiologic response assessments typically encountered in routine clinical care and retrospective datasets; full machine-learning approaches often provide limited interpretability and limited ability to account for temporal dependencies; and available mechanistic models are difficult to parameterise and do not readily account for treatment effects. These limitations complicate the assessment of the temporal dynamics of metastatic spread and motivate the development of a modelling framework that enables interpretable and computationally efficient assessment of metastasis development based on a continuous-time description.

Here, we develop and apply a stochastic disease-progression model based on continuous-time Markov chains (CTMCs). Unlike discrete approaches, the proposed framework jointly models spatial patterns of metastatic involvement and their temporal evolution, enabling estimation of when and where subsequent metastases arise. Transition rates are parameterised to incorporate molecular subtype, histological features, and the modulatory effects of existing metastases on future dissemination. To enable the integration of high-dimensional information on treatment history, we made use of a tailored LLM-based embedding. By applying this model to a large multicentre registry - the Austrian Group for Medical Tumor Therapy (AGMT) registry for metastatic breast cancer (MBC) patients (AGMT MBC registry) - we map both the qualitative pathways and the quantitative speed of progression. Drawing on these findings, we identify specific metastatic profiles associated with a high risk of subsequent cerebral metastasis.

## 2 Results

### A continuous-time Markov chain framework for metastatic disease progression

To analyse metastatic progression trajectories, we developed a disease-progression analysis framework based on AI-assisted continuous-time Markov chains (CTMCs) (Figure 1A,B). This framework enables the analysis of real-world data, where covariate information is highly complex and follow-up as well as radiologic response assessments occur at irregular and patient-specific time points. By operating in continuous time, the framework jointly models the timing and anatomical pattern of metastatic spread while making full use of the information contained in variable observation intervals and heterogeneous patient histories.

**Figure 1:**
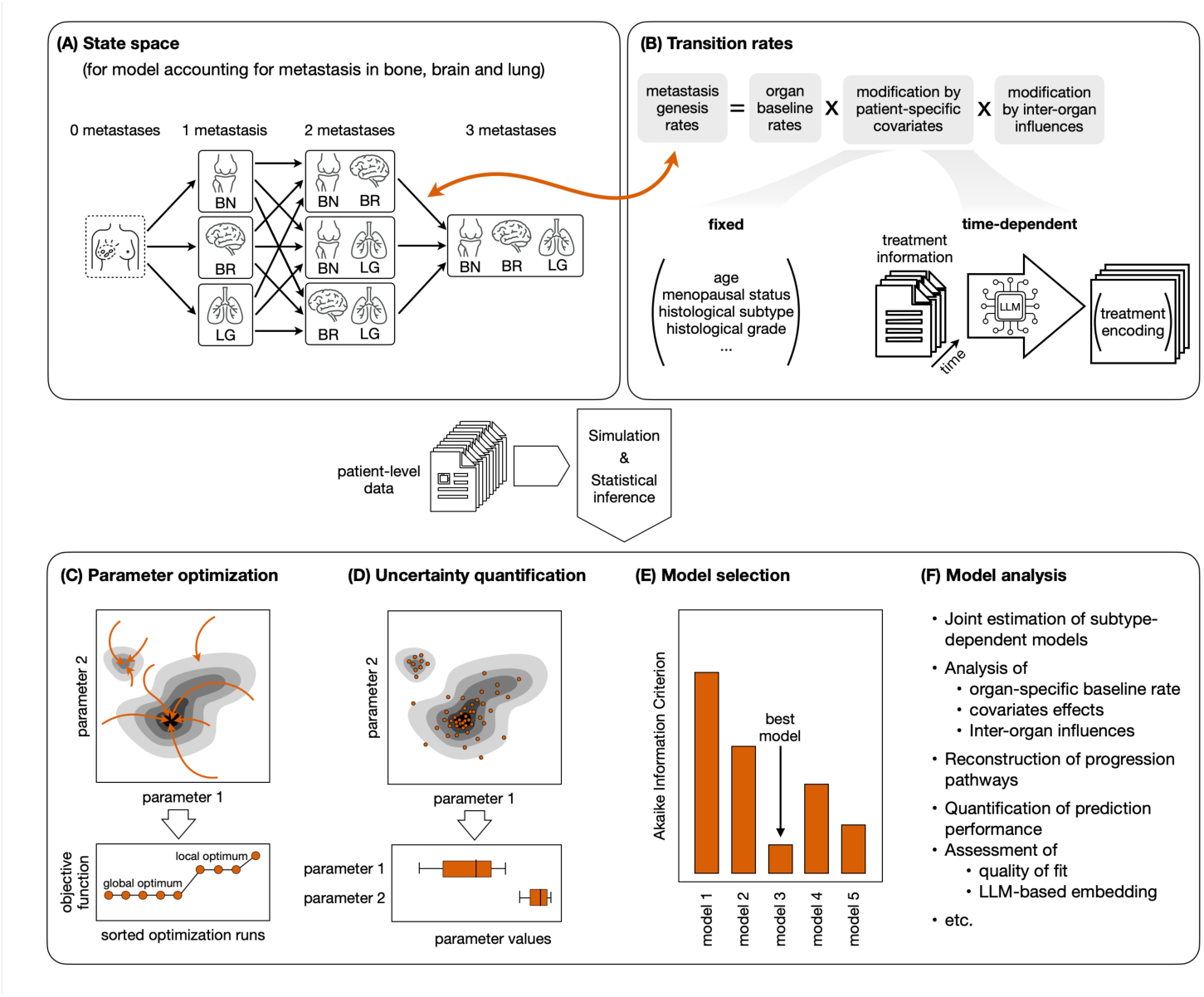
Illustration of AI-assisted continuous-time Markov chain framework for modelling metastatic breast cancer progression. (A) Model states and transitions for a reduced setup with three metastatic locations is shown: bone (BN), brain (BR), and lung (LG). (B) Transition-rate structure, including different types of patient-specific covariates. (C) Maximum a posteriori (MAP) estimates and reliability assessment using waterfall plots. (D) Uncertainty quantification using Hamiltonian Monte Carlo posterior sampling, yielding credible intervals for model parameters. (E) Model selection using the Akaike Information Criterion (AIC). (F) Downstream analyses enabled by the fitted model.

Each patient’s disease state is represented as a configuration of metastatic sites across a set *L* of anatomical categories. This yields a discrete state space of 2^|^*^L^*^|^ possible metastatic configurations, with transitions corresponding to the acquisition of additional metastatic sites. Metastases are assumed to represent persistent disease progression once detected, resulting in a directed, acyclic state topology that reflects biological tumour evolution. This explicit representation of combinatorial metastatic states is necessary to capture co-occurrence patterns and interdependencies between sites that cannot be resolved when modelling sites independently.

Transition rates between states are parameterised in an interpretable and modular manner (Figure 1B), separating three components:

- baseline organ-specific metastasis rates,
- patient-specific covariate effects, and
- inter-organ influence effects that quantify how existing metastases modulate the risk of spread to other organs.

This decomposition enables a disentanglement of intrinsic organ tropism from secondary spread dynamics induced by established metastases, while preserving the direct biological interpretability of all parameters. Importantly, the continuous-time formulation allows inference to exploit the duration between clinical observations, rather than relying solely on the observed ordering of metastatic events, which is often incomplete or ambiguous in intermittently sampled data. Differences between subtypes are accommodated by admitting per-subtype parameters for the baseline rates, inter-organ influences and for selected covariates.

The patient-specific covariates include both static clinical characteristics and treatment-related information. However, treatment histories in metastatic breast cancer are highly heterogeneous and evolve dynamically across therapy lines, such that a naive representation would be high-dimensional and sparse. To incorporate treatment exposure without inflating parameter dimensionality, we summarise treatment characteristics using a pre-trained large language model (LLM) fine-tuned on biomedical data [LSM+21], apply dimensionality reduction to the resulting embeddings, and include the final low-dimensional representation as time-varying covariates in the transition rate model.

The resulting state space and parameterisation introduce substantial computational complexity. To enable scalable inference, we implemented a bespoke likelihood evaluation that exploits the sparsity and acyclic structure of the transition graph, thereby avoiding the need for full matrix exponentiation. Model parameters were estimated using likelihood-based methods (Figure 1C), with uncertainty quantified via Hamiltonian Monte Carlo posterior sampling (Figure 1D), yielding both point estimates and credible intervals for baseline risks, interaction effects, and covariate associations. For the comparison of competing hypotheses, we employed the Akaike Information Criterion (AIC) [Aka98] (Figure 1E).

Together, this framework provides a flexible, scalable, and uncertainty-aware approach for analysing metastatic progression in real-world breast cancer data, enabling downstream inference of subtype-specific interaction patterns and latent progression pathways that are inaccessible to discrete-time or site-independent models (Figure 1F). Mathematical details are provided in the *Materials & Methods* section and Supplementary Text B.

### Model-based analysis of the AGMT metastatic breast cancer registry reveals factors shaping metastatic breast cancer progression

To study the heterogeneity of metastatic breast cancer progression and the spatial distribution of metastases, we used data from our ongoing Austrian Group for Medical Tumor Therapy (AGMT) registry for metastatic breast cancer (MBC) patients (AGMT MBC registry, NIS004886). The prospective and retrospective multicentre AGMT MBC registry provides standardised, highly curated longitudinal documentation of patients with histologically confirmed breast cancer and histological and/or radiological evidence of distant metastases. Tumour characteristics, medical histories, and treatment sequences are documented in anonymised form following written informed consent, with entries for deceased patients permitted without consent.

At the time of analysis, the registry comprised data from 2660 patients with metastatic breast cancer (Figure 2A,B) including 916 luminal-A like, 730 luminal-B like, 306 luminal HER2+, 202 HER2+, and 472 triple-negative breast cancer (TNBC) patients (Figure 2C). The median age at first diagnosis of metastatic disease was 63 years (range 22 – 97, interquartile range 53–74) (Figure 2D). 1048 Patients had a *de novo* mestastatic disease, and among patients with metachronous metastatic disease, the median disease-free survival until first diagnosis of metastatic disease was 45 months (range 0.3 – 400.5, interquartile range 20–90). Across the cohort, 103 distinct systemic treatments were documented, including targeted agents, endocrine therapies, chemotherapies, and immunotherapies, resulting in a large number of observed treatment sequences. The median number of recorded treatment lines per patient was 2 (range 1 – 14, interquartile range 1—4) (Figure 2E). In total, the data included records of 1875 bone, 1282 liver, 1164 lymph node, 1069 lung, and 464 brain metastases (Figure 2F). The metastatic pattern at first diagnosis of metastatic disease was highly heterogeneous (Figure 2G). Patient trajectories also differed substantially in their dynamics and the appearance of new metastases (Figure 2H), with notable dependence on molecular subtype (Supplementary Figure S2). A descriptive overview of the patient cohort is provided in Supplementary Table S17, alongside statistical demographic analyses in Supplementary Text A.

**Figure 2:**
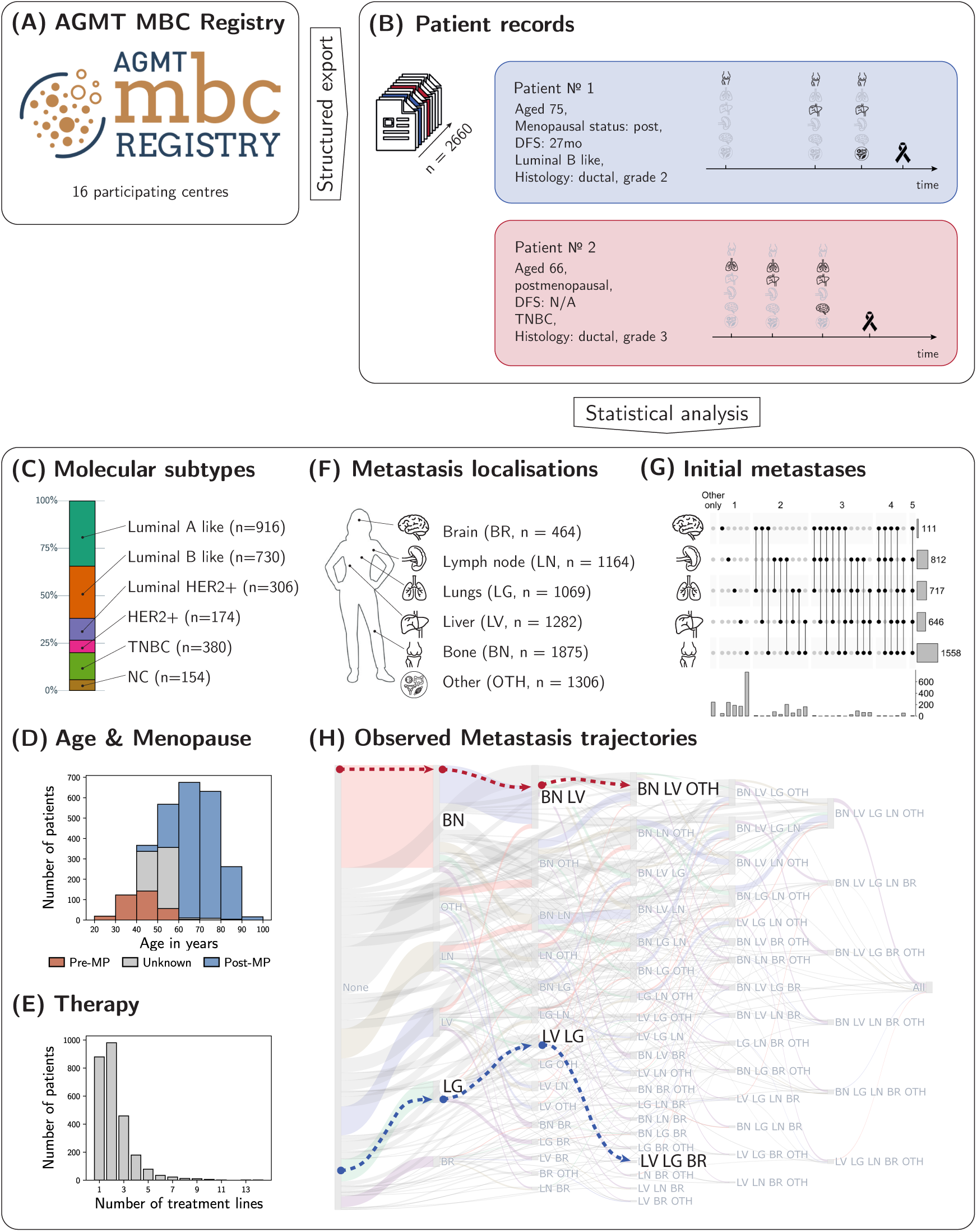
Description of the AGMT MBC registry. (A,B) Visual outline of the assembly and information in the AGMT MBC registry, including two representative patients. (C) Distribution of molecular subtypes. (D) Age and menopausal (MP) status at first diagnosis of metastatic disease. (E) Number of recorded systemic treatment lines per patient. (F) Frequency of documented metastatic sites across the cohort. (G) Metastatic patterns at first diagnosis of metastatic disease. (H) Visualization of the observed data using a Sankey diagram. Individual columns correspond to different numbers of metastases, and individual bars to different combinations of metastases (abbreviations: bone (BN), lung (LG), liver (LV), lymph nodes (LN), brain (BR), and other (OTH)). The transparent areas indicate patient flows, with the colour coding indicating the newly observed metastasis (following the colour scheme in the first transition). As intermediate states are sometimes not observed, flows can skip one or more columns. The height of the bars and flows is proportional to the number of patients. The coloured lines indicate the paths of the representative patients included in (B).

The AGMT MBC registry constitutes a uniquely rich resource for studying metastatic breast cancer progression, combining broad coverage, detailed clinical annotation, and longitudinal follow-up across molecular subtypes. To pinpoint whether patient-specific factors and organ interactions affect metastatic breast cancer progression, we performed model-based hypothesis testing using the proposed framework. We considered a series of nested CTMC models with increasing complexity, starting from models with baseline organ-specific transition rates only and subsequently adding molecular subtype dependence, patient-specific covariates, inter-organ interaction effects, and therapy information embedded as time-varying covariates. These models consider as possible metastasis locations bone (BN), lung (LG), liver (LV), distant lymph nodes (LN), brain (BR), and an aggregated other (OTH) category, yield a discrete state space of 2^6^ possible configurations.

Model selection using AIC consistently favoured the inclusion of molecular subtype dependence (ΔAIC = 560.68, see also Figure 3). Further substantial improvements in AIC scores were observed upon integrating patient-specific covariate effects (ΔAIC = 1141.01) and organ interactions (ΔAIC = 430.78). Incorporating therapy information as LLM-based low-dimensional embeddings further improved model fit (ΔAIC = 119.63), indicating that treatment effects can be captured in a realistic yet parsimonious manner without introducing a high-dimensional representation that would substantially increase the risk of overfitting.

**Figure 3:**
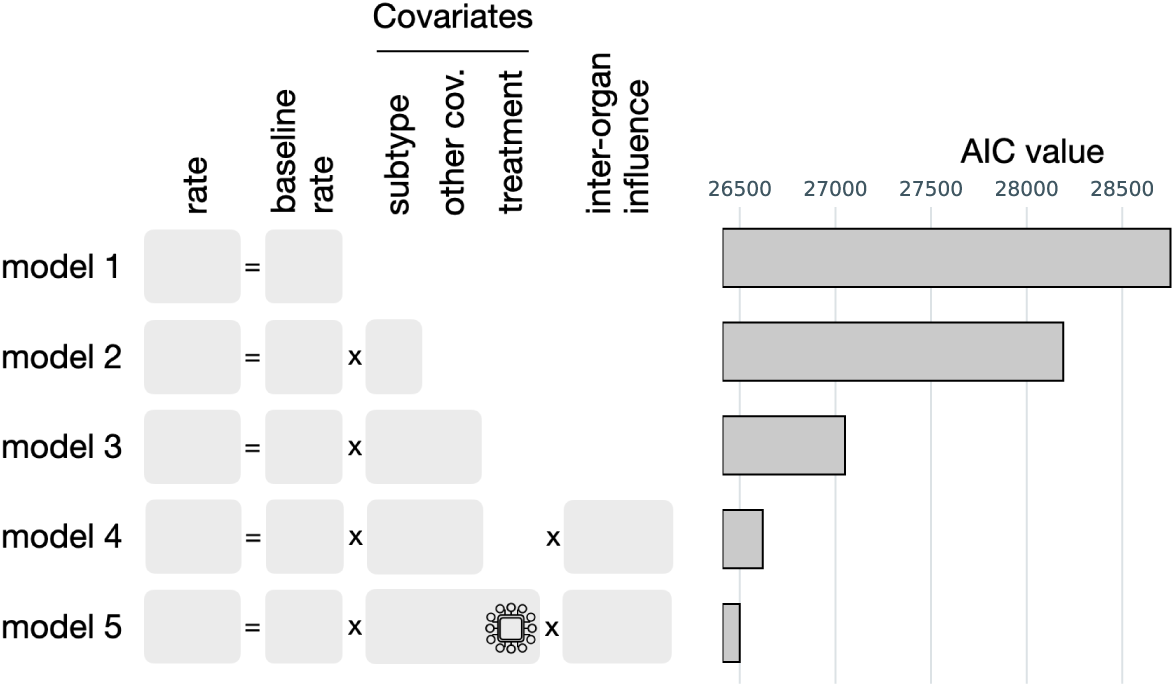
Illustration of model selection and inspection. Model selection via the Akaike Information Criterion (AIC) among models of different complexity levels. Lower values indicate a better score. Model 1 consists only of organ-specific baseline genesis rates. Model 2 makes rates subtype-dependent. Model 3 adds modulation by fixed covariates. Model 4 adds inter-organ influences. Model 5 adds LLM-based treatment effects.

The analysis demonstrates the capacity of the framework to compare different hypotheses and the pinpoint causes of heterogeneity in metastatic breast cancer progresses. Specifically, provides statistical evidence for the importance of both patient-specific characteristics and dependencies between metastatic sites. In the following, we will study the individual contributions in more details.

### Molecular subtypes exhibit substantially different organ tropism

Having selected the model structure, we first quantified how the baseline rates of metastasis formation in different organs differ between molecular subtypes. To this end, we performed Bayesian uncertainty quantification of the selected CTMC model following maximum a posteriori (MAP) optimisation. This yielded posterior estimates and credible intervals for subtype- and organ-specific baseline transition rates, as well as for pairwise contrasts between molecular subtypes.

The inferred baseline rates differed markedly across organs and subtypes. Estimates showed that baseline emergence rates of metastases can vary by as much as an order of magnitude between subtypes. For example, the luminal subtypes exhibited comparable rates of bone metastasis emergence, which were substantially higher than those for the HER2+ and triple-negative subtypes (e.g., *luminal A like* vs. *TNBC* log hazard ratio [95% credible interval] 1.16; 95% CI [0.17, 2.19]). Conversely, luminal A like and luminal B like tumours showed very low rates of de novo brain metastasis compared with HER2+ disease (e.g., *luminal A like* vs. *HER2+* log_2_HR -3.04; 95% CI [-4.26, -1.75]). Subtype-specific differences were observed across multiple organs, as visualised in Figure 4 in terms of both absolute transition rates and pairwise subtype contrasts.

**Figure 4:**
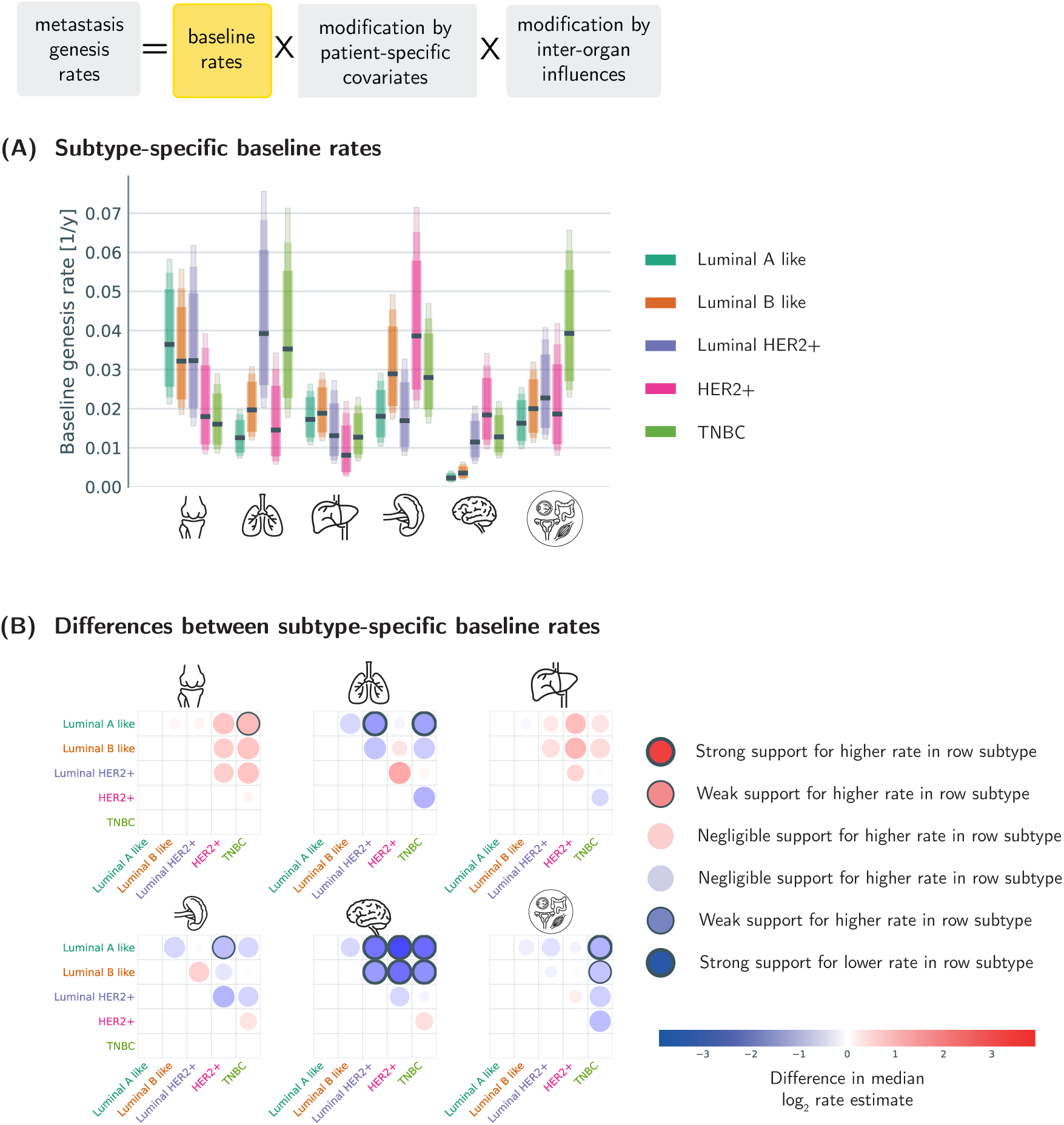
Site-specific baseline rates of metastatic onset by subtype. (A) Baseline rates of metastasis formation by organ and subtype. The black line indicated the posterior median estimates, and the shaded areas the 80% 90% and 95% credible interval. (B) Pairwise subtype contrasts, with bubble colour corresponding to magnitude of difference (row subtype minus column subtype), and bold borders indicating contrasts whose 95% and 99% credible intervals, respectively, do not cover zero.

These findings identify molecular subtype as a major determinant of organ-specific metastatic risk, resulting in different organ tropism. The estimated rates recapitulate established subtype-dependent patterns of organ tropism, including preferential bone involvement in luminal disease and increased propensity for cerebral metastasis in HER2+ disease.

### Patient-level covariates explain additional heterogeneity in metastatic progression

After assessing subtype- and organ-specific baseline rates, we studied the effects of additional patient-specific covariates using the posterior distribution of the selected model. Covariates were organised into shared covariates, which exert the same effect across molecular subtypes, and subtype-specific covariates. Overall, we considered tumour grade, Ki-67 status, histological subtype, disease-free survival, age at first diagnosis of metastatic disease, menopausal status, treatment line, and embedded treatment information (Figure 5).

**Figure 5:**
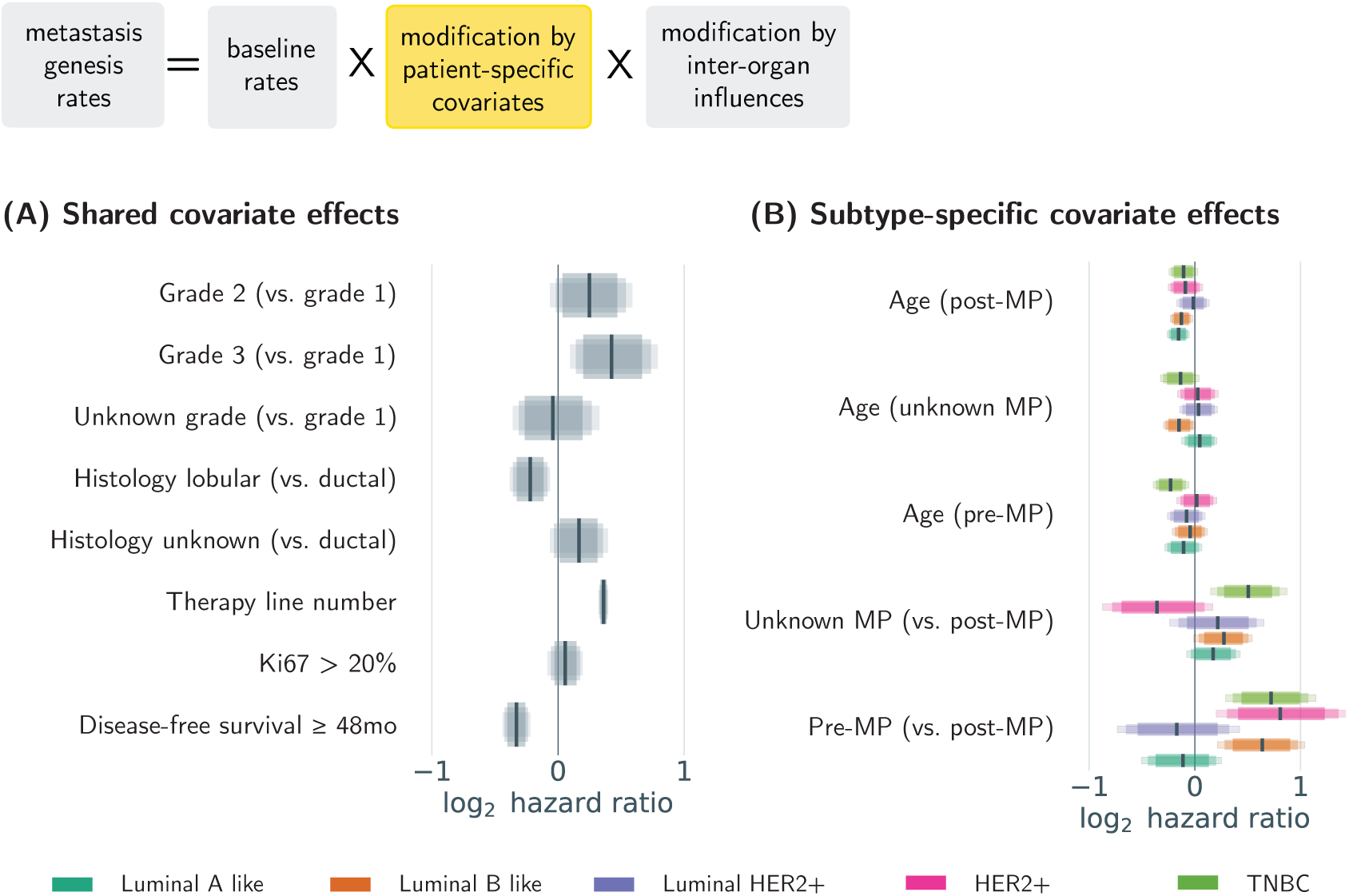
Patient characteristics associated with metastatic progression across organ sites. Estimated covariate effects on metastatic transition rates, modelled on a log-linear scale. Values greater than 1 indicate accelerated progression; values less than 1 indicate reduced progression. Horizontal bars represent 95% credible intervals. Panel A shows estimated effects of covariates shared across subtypes; panel B shows subtype-specific effects of the demographic covariates age and menopausal (MP) status.

The highest estimated effects among the shared covariates were observed for tumour grade and number of treatment lines (Figure 5A). Higher tumour grade was associated with increased metastatic progression risk (grade 2 vs. 1 log_2_HR 0.25; 95% CI [-0.06, 0.59]; grade vs. 1 log_2_HR 0.42; 95% CI [0.10, 0.79]), and each treatment line beyond the first was associated with a higher transition rate (log_2_HR 0.36; 95% CI [0.32, 0.40]). Elevated Ki-67 showed a weaker association with progression risk (log_2_HR 0.06; 95% CI [-0.08, 0.20]). In contrast, long disease-free survival (log_2_HR -0.33; 95% CI [-0.44, -0.22]) and lobular-spectrum histology (log_2_HR vs. ductal/NST -0.22; 95% CI [-0.38, -0.06]) were associated with lower progression rates.

The estimated effects for the subtype-specific covariates confirmed the assumed subtype dependence. Pre-menopausal status, as compared to post-menopausal, was associated with an increased risk of progression in luminal B like (log_2_HR 0.64; 95% CI [0.21, 1.04]), HER2+ (log_2_HR 0.81; 95% CI [0.20, 1.46]), and TNBC (log_2_HR 0.72; 95% CI [0.29, 1.15]). Higher age (measured as decades from 60 years) at first diagnosis of metastatic disease was associated with lower progression rates in several patient groups, namely luminal A like post-menopausal (log_2_HR -0.16; 95% CI [-0.26, -0.05]), luminal B like post-menopausal (log_2_HR -0.13; 95% CI [-0.23, -0.02]), and TNBC pre-menopausal (log_2_HR -0.23; 95% CI [-0.39, -0.05]).

With respect to the treatment embeddings, visual inspection showed agreement between the embedding regions and pharmacological groups. The estimation resulted in mostly limited estimated effects (see Figure 6 for a visualisation of the first two component results, as well as Supplementary Figure S3 for an overview over all embedding dimensions). However, individual embedding vectors can and should not be interpreted directly, as therapy decisions are inextricably linked to patient state, leading to implicit bias which cannot be accounted for.

**Figure 6:**
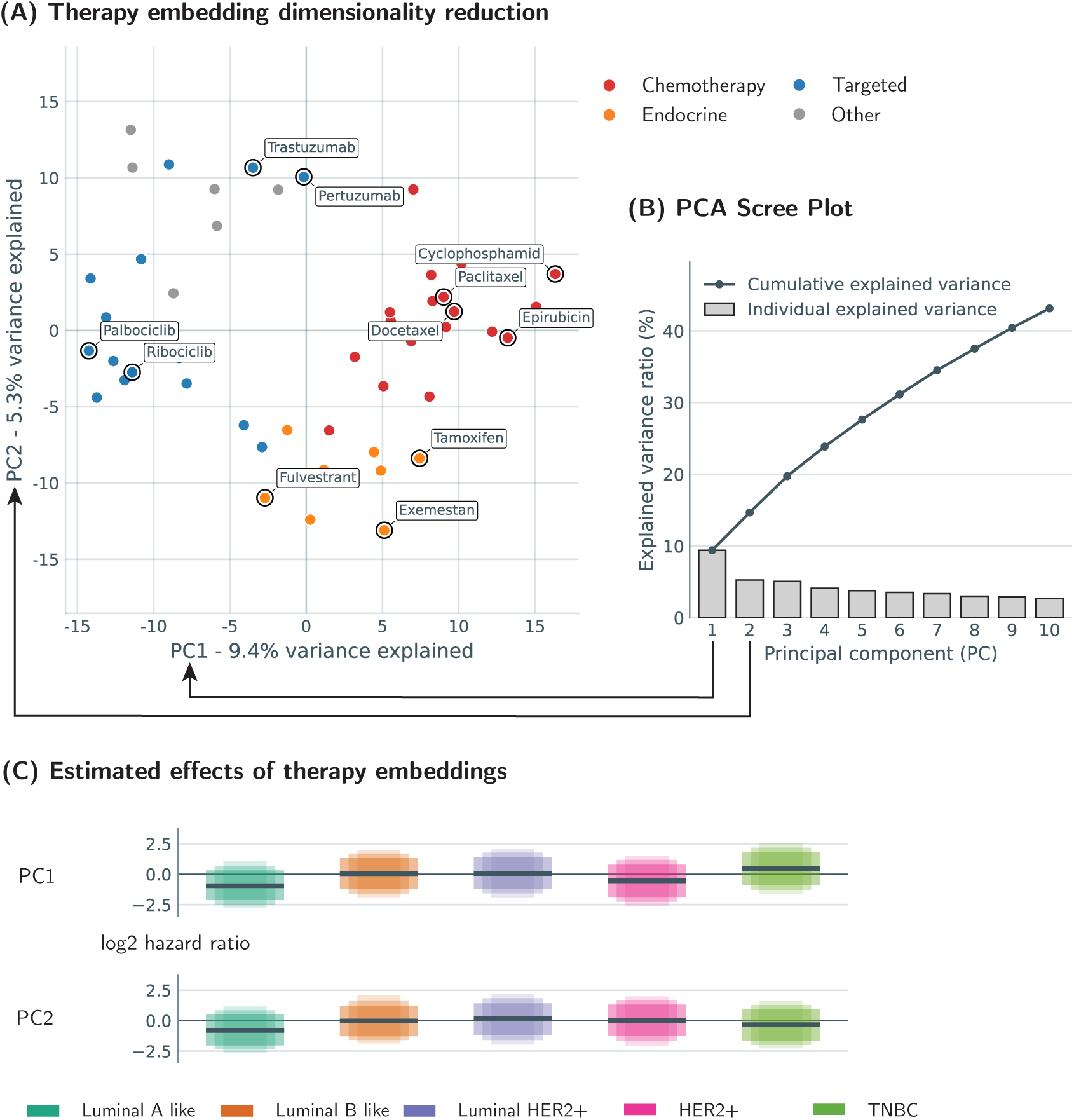
Analysis of LLM-embedded treatment effects. Visualisation of treatment embeddings and associated effects. Panel A shows a scatter plot of the first two principal components. Each dot represents a therapy, and its position is determined by projection of the 768-dimensional LLM embedding onto the two directions of highest variance. In panel B, the individual principal component explained variance ratios are shown, as well as the cumulative explained variance upon considering the first *n* components. In panel C, the estimated log-linear effects on progression by the respective treatment PCs are shown. The black line indicates the posterior median estimates, and the shaded areas the 80% 90% and 95% credible interval.

### Inter-organ interaction estimates reveal subtype-specific progression patterns

Having quantified baseline organ tropism a1n4d patient-level covariate effects, we assessed whether metastatic progression is further shaped by dependencies between anatomical sites. To this end, we considered the model parameters encoding how the risk of metastasis formation in one organ depends on the presence of established metastases in other organs. As these interaction effects were estimated jointly while adjusting for subtype and patient-level covariates, these associations cannot be attributed to marginal correlations or limited stratified analyses, but instead reflect consistent structure across the full dataset.

A number of interactions was found to be supported by the statistical evidence (see Figure 7). In total, for 41 interactions (across the five subtypes) the estimated posterior distributions’s 95% credible interval did not cover zero, suggesting at least weak support for an interaction. Of these, 23 had a posterior distribution whose 99% credible interval did not cover zero, indicating strong support for an interaction. Where effects were clear, existing metastases were overwhelmingly associated with accelerated progression. The strongest effects were observed to be lung on liver in TNBC (log_2_HR 2.02; 95% CI [1.26, 2.75]), liver on bone in TNBC (log_2_HR 1.77; 95% CI [0.98, 2.51]), lung on liver in HER2+ disease (log_2_HR 1.70; 95% CI [0.42, 3.00]), Other on lung in HER2+ disease (log_2_HR 1.70; 95% CI [0.49, 2.91]), and bone on liver in luminal A like disease (log_2_HR 1.67; 95% CI [1.17, 2.28]). Some interactions were found to be present across all subtypes; such as the influence of liver on brain metastases (luminal A like log_2_HR 1.16; 95% CI [0.48, 1.87], luminal B like log_2_HR 1.18; 95% CI [0.48, 1.89], luminal HER2+ log_2_HR 1.11; 95% CI [0.50, 1.76], HER2+ log_2_HR 0.75; 95% CI [-0.03, 1.51], TNBC log_2_HR 0.86; 95% CI [0.19, 1.52]). Others were present across the luminal like subtypes, but not the others, such as the influence of bone on liver metastases, (luminal A like log_2_HR 1.67; 95% CI [1.17, 2.28], luminal B like log_2_HR 1.11; 95% CI [0.61, 1.63], luminal HER2+ log_2_HR 1.34; 95% CI [0.32, 2.32], HER2+ log_2_HR 0.32; 95% CI [-1.03, 1.75], TNBC log_2_HR 0.37; 95% CI [-0.41, 1.09]) – or vice versa, such as the influence of lung on bone metastases (luminal A like log_2_HR 0.05; 95% CI [-0.57, 0.66], luminal B like log_2_HR 0.42; 95% CI [-0.24, 1.04], luminal HER2+ log_2_HR 0.07; 95% CI [-1.01, 1.03], HER2+ log_2_HR 1.29; 95% CI [0.19, 2.28], TNBC log_2_HR 0.94; 95% CI [0.21, 1.67]). Yet other estimated influences were specific to a subtype, such as the contribution of Other-localised metastases to lung metastasis hazard in the HER2+ subtype (HER2+ log_2_HR 1.70; 95% CI [0.49, 2.91]; luminal A like log_2_HR 0.18; 95% CI [-0.43, 0.81], luminal B like log_2_HR 0.38; 95% CI [-0.15, 0.94], luminal HER2+ log_2_HR 0.15; 95% CI [-0.76, 1.06], TNBC log_2_HR -0.34; 95% CI [-1.22, 0.48]).

**Figure 7:**
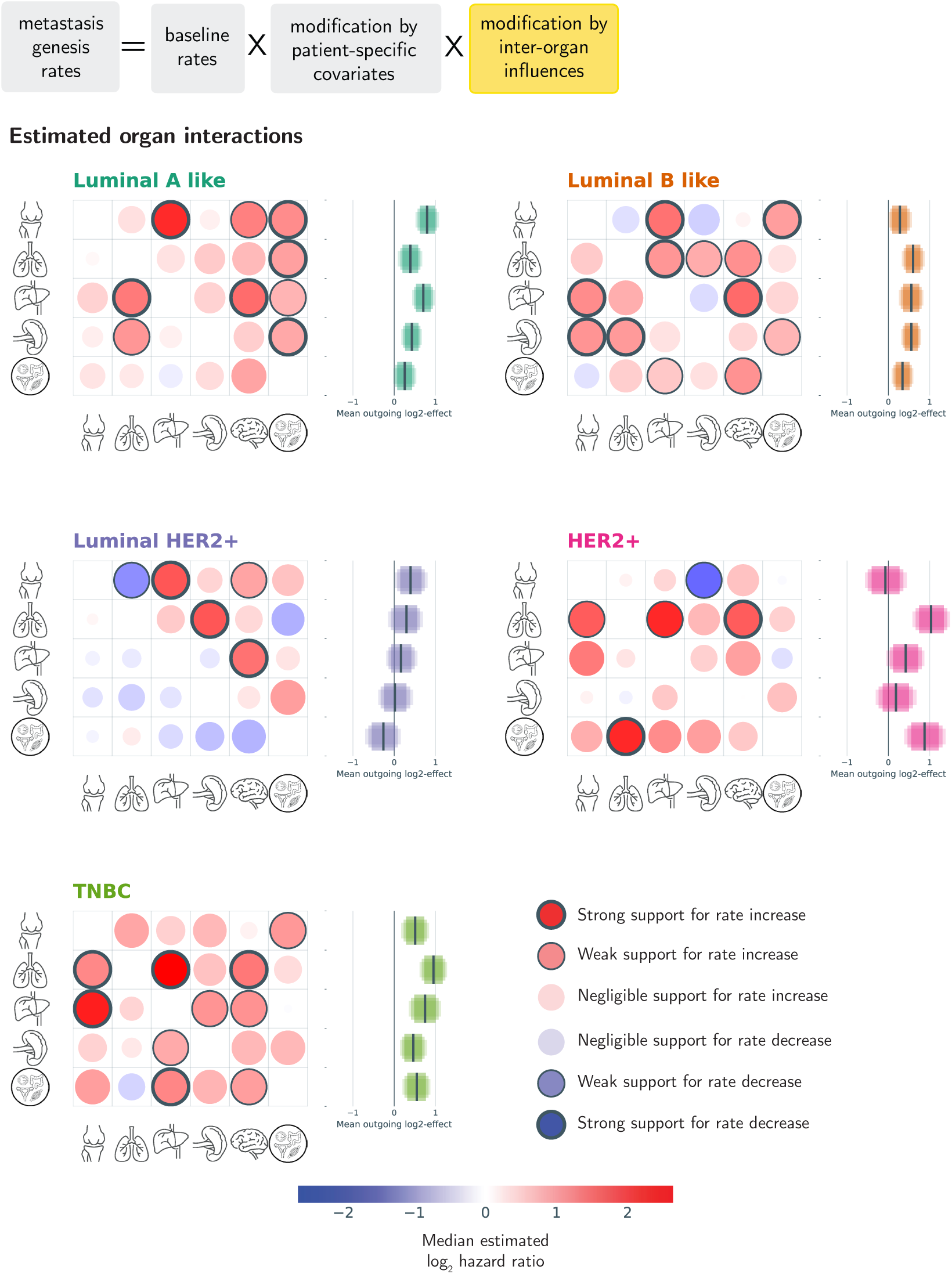
Organ interactions driving metastatic progression. Bubble plot showing the estimated strength of interactions between metastatic sites, with rows showing representing source, and columns target organs. Bubble colour reflects the direction and magnitude of the interaction effect; bubble size represents the confidence that the effect deviates from zero, based on the posterior distribution. Interactions whose 95% and 99%, respectively, credible intervals exclude zero are highlighted with bold outlines.

To identify which metastatic sites act as major accelerators of disease progression, we quantified the overall increase in metastatic risk associated with the presence of each organ-specific metastasis. Using the inferred transition rate parameters and their uncertainties, we computed subtype-specific aggregate risk increases attributable to each metastatic site. While this is in line with the observed risk increase of later treatment lines, comparing its magnitude to the aggregate increases in progression risk reveals that in many cases, existing metastases exert a much stronger effect on further progression. The estimated effect of an additional treatment line (log_2_HR 0.36; 95% CI [0.32, 0.40]) is much smaller than many of the aggregated effects on progression of established metastases, such as lung metastases in the HER2+ subtype (1.03; 95% CI [0.58, 1.49]), lung metastases in TNBC (0.95; 95% CI [0.62, 1.28]), Other sites in HER2+ (0.88; 95% CI [0.38, 1.38]), bone in luminal A like (0.80; 95% CI [0.52, 1.08]), or liver in TNBC (0.75; 95% CI [0.35, 1.14]).

Only two instances of interactions supported by the statistical evidence at the 5% level carried a negative sign – the bone to lung influence in luminal HER2+ (log_2_HR -0.89; 95% CI [-1.72, -0.04]) and bone to lymph node influence in HER2+ (log_2_HR -1.19; 95% CI [-2.40, -0.09]). In both cases, however, this was accompanied by estimated increases to other organ metastasis rates, meaning that the overall effect is not protective, but rather simply re-orienting the subsequent progression distribution.

Overall, the interaction estimates show that metastatic sites cannot be treated as independent events. Instead, established metastases are associated with subtype-specific changes in subsequent dissemination risk, supporting the need for a model structure that explicitly accounts for inter-organ dependencies.

Overall, the AGMT MBC registry enables well-determined parameter estimates for a broad range of clinically relevant covariate effects, reflecting the size and depth of the longitudinal dataset. At the same time, several estimates remain uncertain, particularly for subtype-specific and treatment effects involving small patient subgroups or rare metastatic progression patterns.

## 3 Discussion

In this study, we present an analysis framework for stochastic modelling of disease progression in metastatic breast cancer, which we use to analyse the AGMT MBC registry. This approach leverages the flexibility and deep mathematical theory of discrete-state Markov processes, making it well-suited for the analysis of longitudinal real-world data, where staging assessments are performed at variable, practice-driven time points. Equipped with a biologically informed parametrisation and formal uncertainty quantification, it captures both patient-specific risk factors and the influence of established metastases on subsequent dissemination. This enables to capture the considerable heterogeneity in patient trajectories and outcomes, while providing the possibility of deriving concrete biological associations from the resulting estimates. Applying this method, we identify both established and previously unrecognised patterns governing metastatic spread.

At a covariate level, the increase in metastasis rate for patients of higher histological grade [GWH+22] and the prognostic value of disease-free survival [KAL+15; CMA+19; SGZ+17], where a longer metastasis-free interval has been described as predictive of a more indolent course, are consistent with previous reports. The much less clear impact of Ki-67 status, too, reflects differences in the literature. [ACC+07; NLR+21; LNV+23].

Investigating the relationship between breast cancer subtypes and patient trajectories, Xiao et al. [XZY+18] have previously described the association of HR+/HER2+ and HR-/HER2+ subtypes with higher rates of liver, brain, and lung metastases. They have furthermore reported TNBC patients to have a higher rate of brain, liver and lung metastases, but a lower rate of bone metastases compared to HR+/HER2- tumours. Some of these findings are directly mirrored by the baseline rate estimates obtained in our study; especially the higher tendency towards brain metastases in HER2+ and TNBC, and conversely the lower rate of bone metastasis in TNBC. While we do not estimate the HER2+ subtype to confer an increased baseline risk of visceral metastasis, our findings remain consistent with these previous observations when taking into account the strong interaction effects, e.g. the acceleration of lung metastasis in the HER2+ subtype by established Other-localised metastases (log_2_HR 1.70; 95% CI [0.49, 2.91]).

Similarly, there is broad agreement on the increased risk of brain metastasis in patients with HER2+ or TNBC, with substantially higher incidence reported by Kuksis et al. [KGT+21]. Transforming them – given as 0.13 per patient-year for the HER2+ subgroup, and only 0.05 for patients with HR+/HER2- MBC – into an incidence ratio of approximately 0.37, the effect size is comparable to the rate ratios we estimate, with a disaggregation into luminal A like 0.12; 95% CI [0.05, 0.30], and luminal B like 0.19; 95% CI [0.08, 0.45].

Beyond these established associations, our analysis provides quantitative evidence for inter-organ dependencies in the metastatic progression, resolved at the subtype and organ levels. These findings support the concept that metastatic spread is not a sequence of independent events, but rather a dynamically coupled process shaped by interactions between tumour sites. The individual elements of our findings are in concordance with previous work. For example, Zhang et al. have previously described the role of the bone micro-environment in priming tumour cells for secondary metastasis [ZBH+21] in luminal and triple-negative breast cancer, contributing to the observation by Huang et al. that “bone serves as a transfer station for secondary dissemination of breast cancer” [HWY+23]. Our holistic modelling approach extends this insights by quantifying the impact of established bone metastases on subsequent organ-specific spread, and by identifying which downstream sites are most strongly affected, such as the preferential association between bone and liver involvement in Luminal A disease.

Brain metastases warrant particular attention due to their clinical impact and associated morbidity. Previous studies have contributed evidence that visceral [WNR+22], and in particular lung [BBD+12], metastases are associated with an increase risk for central nervous system metastasis. Our findings extend these observations by demonstrating subtype-specific differences in these risk patterns, with lung metastases playing a more prominent role in HER2-positive and triple-negative disease, and liver metastases contributing more strongly in luminal subtypes. In addition, we identify a compounding effect of bone metastases in luminal A like and luminal HER2+ disease. In a general breast cancer cohort, Kim et al. [KKK18] report an overall brain metastasis incidence for the HER2+ subtype of 0.01, but note that among HER2+ patients with multiple extracranial (specifically bone, liver and lung) metastases, the incidence per patient-year rises as high as 0.28. This is comparable with our estimates of interaction effects in the HER2+ subtype: Influences on brain metastases respectively by bone (log_2_HR 0.49; 95% CI [-0.31, 1.26]), liver (log_2_HR 0.75; 95% CI [-0.03, 1.51]), and lung metastases (log_2_HR 1.29; 95% CI [0.48, 2.04]), together with the metastasis risk effects attributable to additional treatment lines upon progression, imply a 12-fold increase in rate; showing how our methodological framework can be applied to disentangle intertwined effects. These results directly address a clinically unmet need for personalised risk assessment of brain metastases. Current guidelines recommend screening primarily in in the presence of neurological symptoms [GAB+21; KAF+13; DUK+20; GMA+23]. However, a recent survey by Matos et al. [MRM+25] has found that indeed 35% of physicians never perform screening for brain metastases under any circumstances, and only 13% routinely address the issue of cerebral metastasis routinely with their patients – while 91% of patients would like to receive information on this topic. This uniform, symptom-triggered approach has been criticised for delayed detection and suboptimal outcomes [CMC+18], particularly when compare to other cancer entities such as non-small cell lung cancer (NSCLC). Our findings provide a data-driven basis for more individualised surveillance strategies, which could enable earlier detection and potentially less aggressive management [TON+23; KWM+20; AKA+25].

Methodologically, our work can be seen as a natural extension of previously proposed continuous-time multi-state models of disease progression in a number of tumour entities, which have already demonstrated the value of properly uncertainty-aware probabilistic modelling. Previous studies, however, were limited to coarse state spaces. For example, Farahani et al. [FDR+19] modelled breast cancer progression using only four states: *Primary treatment*, *Recovery*, *Metastasis*, and *Death*. Similarly, Nicora et al. [NMS+20], in the context of myelodysplastic syndromes, demonstrated a continuous-time Markov Chain with a state space with five ordered progression states, spanning from *very low* to *very high*. On the other hand, existing models with individual organ-level resolution, have largely been restricted to discrete-time Markov chain approaches, which discard information on the exact timing between staging assessments. Such approaches have been demonstrated by Newton et al. for lung [NMB+12] and breast [NMV+15] cancer. Our framework addresses this limitation by enabling continuous-time modelling in a high-dimensional state space, preserving both temporal and spatial resolution — an approach that is, to our knowledge, novel. Moreover, the utilisation of a foundation LLM trained on a biochemical corpus gives us the possibility to express longitudinal treatment information in terms of compact embeddings, and thus restrain the number of free estimable parameters.

Comparing our study to that of Newton et al., which classify metastatic sites as “spreaders” or “sponges”, our findings suggest a predominantly facilitatory pattern of inter-organ interactions. For example, liver metastases, categorised by Newton et al. as a “sponge” across subtypes, are consistently associated with an increased progression hazard in our model. In contrast to the work of Newton et al., in which about half the localisations are classified as sponges, we do not observe a clear protective effect of specific metastatic sites. This can have a variety of structural reasons; first and foremost however we hypothesise that it is the consequence of including inter-observation times, which were previously omitted, into our model.

Several limitations should be considered. First, genomic and mutational data are not available in our dataset. For instance, Nguyen et al. [NFL+22], have reported that specific mutational signatures and genomic alterations acquired early in tumourigenesis influence the propensity and timing for metastatic spread to different organs. However, the proposed framework is fully compatible with such genomic data and can be extended in future studies. Second, the “other” metastasis category aggregates heterogeneous sites, potentially obscuring organ-specific biological effects. It is made necessary by the high variety in metastasis localisations, but by necessity leads to a pooling of biologically very different organ systems. In future studies including larger patient populations, this could be disaggregated further, by admitting additional organs with sufficient prevalence in the study population as separate localisations into the model. Third, our analysis is restricted to patients with established metastasic disease, and therefore cannot be generalised to the primary breast cancer population, as this would introduce substantial selection bias related to tumour biology and metastatic potential. Lastly, a note of caution should be added since our modelling approach does not directly distinguish between associations and true causative effects. This would require the inclusion of even larger data pools to enable modelling so fine-grained as to make precise counterfactual simulation possible.

Despite these limitations, our study has several key strengths. The richness and depth of the AGMT MBC registry, which captures longitudinal, temporally resolved patient trajectories at the level of individual organ systems, combined with a novel modelling framework that enables inference on a high-resolution state space, allow for a substantially more detailed characterisation of the effects driving metastatic progression.

Importantly, several of the organ-specific interactions identified in this study, such as the pre-eminent role of lung metastases in HR- subtypes, would not have been detectable without this combination of high-quality real-world data and advanced methodology. In addition, our work further provides a unifying context to interpret broader patterns of disease progression. For example, it highlights how the previously reported increase in metastasis risk of later treatment lines can be re-interpreted in light of specific impacts of established metastases on further progression.

Further research is needed to obtain validated consensus findings that can be translated into actionable recommendations. First and foremost, we believe that leveraging more sources of data is the path towards obtaining more robust results. This can be done through assimilation of un-curated real-world data into existing resources [PPE+25], or data pooling and integration within and between institutions and consortia. Where data sharing is ruled out by privacy concerns, promising solutions include federated learning, a field which allows for decentralised machine learning and modelling, drawing on many datasets while preserving anonymity [RHL+20].

The need for more data goes beyond merely increasing sample sizes. It also encompasses “richer” data, which includes further complementary modalities, to provide an ever more holistic understanding of the processes influencing disease progression. In this sense, our study demonstrates a way for integrating further information, such as imaging data, by utilising foundation models – themselves a field of intense scholarly activity – for adaptation into biologically interpretable analysis.

To conclude, we have presented a comprehensive investigation into the driving dynamics of metastatic breast cancer progression; supported by high-quality data from a large patient population, a flexible stochastic modelling framework incorporating foundation model treatment embeddings, and an implementation tailored to a metastasis configuration state space. Our analysis reveals strong evidence for intricate and subtype-specific inter-dependencies among metastatic sites, hinting at features of underlying biology. At the same time, they agree with existing literature on known clinical risk factors, supporting their credibility. The proposed methodology provides novel possibilities for identifying future criteria to guide screening decision-making, and contributes towards the advancement of personalised medicine.

## 4 Materials & Methods

### 4.1 Study design and outcome parameters

#### Patients

The AGMT MBC Registry is an ongoing, multicenter registry for MBC patients in Austria. For this analysis, patients with available hormone receptor (HR) and HER2 status, as well as sufficient survival data, were included.

When multiple tumor samples were available per patient, a predefined hierarchy was applied. Preference was given to biopsies from a metastatic site obtained within 3 months of the diagnosis of metastatic disease, provided that at least estrogen receptor (ER) and HER2 status were available; in these cases, receptor status, grade, Ki-67, and histologic subtype were derived from this sample. If no such metastatic biopsy was available, data from the most recent primary tumor or local recurrence diagnosed before — or within 3 months of — the diagnosis of metastatic disease were used instead.

Breast cancer subtypes were defined based on ER status, HER2 status, histologic grade, and Ki-67 proliferation index. ER-positiv tumors and negative HER2 status were classified as luminal-like and further subdivided into luminal B–like (grade 3, or grade 2/unknown with Ki-67 ¿ 20%) and luminal A–like (all remaining luminal-like tumors not meeting the luminal B–like criteria). Tumors with concomitant ER and HER2 positivity were classified as luminal HER2+, ER-negative/HER2-positive tumors as HER2+, and ER-/HER2-negative tumors as triple-negative breast cancer (TNBC).

For a visualisation of the overall observed progression trajectories, see Supplementary Figure S2.

#### Outcomes

At the time of diagnosis of metastatic disease and at each subsequent therapy change due to disease progression, the presence and localization of metastatic lesions were systematically documented. Metastatic sites were determined based on routine clinical imaging assessments, primarily contrast-enhanced computed tomography scans of the thorax, abdomen, and pelvis, as well as on clinical evaluation where applicable, such as in patients with cutaneous metastases.

### 4.2 Statistical demographic analysis

The descriptive patient characteristics were compared by using omnibus Kruskal-Wallis tests for continuous, and *χ*^2^ tests of independence for categorical variables. For further pairwise comparisons, Mann-Whitney tests were used for continuous, and *χ*^2^ independence or Fisher exact tests for categorical variables. Bonferroni correction for multiple testing was applied.

### 4.3 Model-based analysis framework

To analyse patients’ disease progression in terms of metastatic sites, we employed a Continuous-Time Markov Chain (CTMC). This is a stochastic model which operates on so-called *states*, which we take to be all possible configurations of metastases. Let *L* be a set of metastatic sites, and *N* = |*L*|. Then, the state space consists of the 2*^N^* possible configurations of metastases, and the proposed CTMC expresses the probability of a *transition* (e.g., that a patient with only a bone metastasis is found to have bone, liver, and brain metastases one year later) given *transition rates* – the key quantities governing its behaviour. Given a set *S* ⊆ *L* of metastases (e.g. {Bone}), we consider the rate of progressing into a new state *S*^′^ (for example {Bone, Liver}) by adding an organ *O*^′^ (in the exemplary case Liver). We choose the transition rates to consist of three factors

1. A subtype-specific *baseline genesis rate* with which metastatic seeding to *O*^′^ occurs;
2. Modification accounting for the patient’s *covariate values* (e.g. demographic or histological information, but also therapy), denoted *x*; and
3. Modification accounting for inter-organ influences. These (learned) subtype-specific coefficients determine how an established metastasis in one organ changes the risk in another (in the example case, a modulating effect by the existing bone metastasis on the genesis of a new liver metastasis)

Specifically, if the new *S*^′^ = *S* ∪ {*O*^′^}, and covariate values are given by *x*, we consider the ansatz

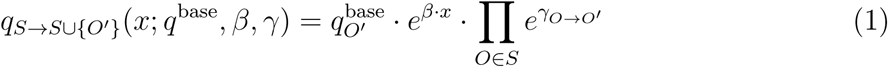

where the *q*^base^, *β* and *γ*, are the model’s degrees of freedom. This formulation gives rise to a likelihood function: Let *i, j* be two states, *ϑ* represent (*q*^base^*, β, γ*, and *S_ij_* be the set of all strictly increasing sequences of integers starting at *i* and ending at *j*. Let furthermore the (*q_ii_*)*_i_* be pairwise distinct. Finally, let in the following *q_ij_* represent *q_i_*_→_*_j_*(*x*; *ϑ*) Then,

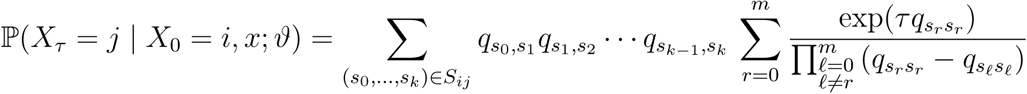

A detailed mathematical account is given in Supplementary Section B.

This likelihood function can now be utilised for Bayesian inference, combining with priors on the model variables to yield an un-normalised posterior density. Knowledge gain from such modelling is now achieved within a Bayesian framework through assessing the estimated posterior distributions, as the model’s free parameters *q*^base^, *β*, and *γ* all represent informative quantities. The *q*^base^ directly give metastasis genesis risks, the *β* parameters represent the modulation of progression risk by covariates, and the *γ* coefficients indicate the influence of established metastases on the progression process.

For the analysis in this study, we consider metastases at the following localisations: Bone, Lungs, Liver, Lymph nodes, and Brain. All further localisations are summarised as an “Other” meta-localisation, yielding a total of six modelled metastatic sites and 64 different possible configurations. All pairwise inter-organ influences were admitted with the exception of those originating at brain metastases, due to the high mortality associated with them. Baseline genesis rates and inter-organ influences were modelled separately per subtype.

### 4.4 Fixed effect and LLM-based treatment encoding

The covariates admitted into the model were chosen as

- Age at first diagnosis of metastatic disease,
- menopausal status at first diagnosis of metastatic disease,
- *disease-free survival* duration (DFS, defined as the time between first primary diagnosis and first diagnosis of metastatic disease),
- the number of prior treatment lines,
- Ki-67 at first diagnosis of metastatic disease,
- histological subtype, and
- histological grade.

Ki-67 status was dichotomised with a threshold at 20%. Menopausal status was admitted as a categorical covariate, with a subtype interaction, effectively estimating separate coefficients per subtype. Age was included as a continuous covariate using decades from the age of 60 as units for numerical stability, with a separate slope per subtype and menopausal status category.

Furthermore, we included covariates to account for possible influences of treatment on progression. For this purpose, we employed SapBERT, a pre-trained large language model for biomedical terms published by Liu et al. [LSM+21], to transform patient therapies to a latent space, 768-dimensional as per the SapBERT architecture, which was then reduced to ten dimensions, determined by inspection of the scree plot to cover 75% of variance, using Principal Component Analysis (PCA). These ten condensed PCA features were rescaled to [−1*/*10, 1*/*10] for numerical stability, and admitted as continuous, time-varying, covariates updated at the start of each new treatment line, acting on transition rates with per-subtype effect parameters.

The parameters for the remaining covariates were shared across subtypes. Histological subtype was included as a categorical variable, with distinct levels for ductal / no special type, and lobular / mixed-lobular, while classes not covered by this distinction were grouped into and *Other* category. Similarly, histological grade, and the dichotomised indicator whether disease-free survival was at least 48 months were admitted as categorical covariates. The number of prior treatment lines was admitted as a pseudo-continuous variable.

### 4.5 Implementation

To ensure identifiability as well as numerical stability, we opted for Bayesian 𝒩 (0, ½) priors with on the parameters encoding covariate effects and inter-organ influences, while keeping non-informative flat priors on the remaining parameters. To assess convergence of the sampling procedure, we calculated rank-normalized split-R^ [VGS+21] and the Gelman–Rubin potential scale reduction factor (PSRF) [GR92; BG98].

We implemented the log-posterior density in a way compatible with automatic differentiation (AD), in particular through the `ForwardDiff.jl` framework [RLP16]. As a slow fallback in case of numerical instability, we utilised the AD-compatible matrix exponential implementation provided by `ExponentialUtilities.jl` [RN17]. To obtain the treatment encodings, we accessed the pre-trained `SapBERT` model through the `huggingface` [Jai22] software package which provides wrapper access to serialised PyTorch [PGC+17] transformer models.

Maximum a posteriori point estimates were obtained by using multi-start optimisation with the `BlackBoxOptim.jl` [FS18] toolkit. Furthermore, we calculated as parameter credible intervals by using Hamiltonian Monte-Carlo (HMC) [DKP+87] sampling as implemented by the `AdvancedHMC.jl` [XGT+20] software package. The MCMC convergence checks were carried out using an implementation provided by `MCMCDiagnosticTools.jl` [Tur21]. For detailed information about data processing inference hyper-parameters, and the obtained diagnostic values, see Supplementary Section C.

The analyses were carried out using Python v3.11, and Julia v1.10. All model variants were trained on data from the AGMT MBC registry.

## Data Availability

The clinical data can be shared upon approval of the analysis proposal by the Steering Committee and sponsor of the AGMT MBC registry, and after a data sharing agreement has been signed. Requests for data sharing should be addressed in writing to the scientific CEO of the SCRI-LIMCR GmbH and the AGMT GmbH, i.e. to R.G..

https://github.com/marc-vaisband/MBC-CTMC

## Acknowledgements

The authors gratefully acknowledge the support from the AGMT office and all trial coordinators at the contributing centres.

Generative artificial intelligence was used to support coding. The authors take full responsibility for the correctness of the code provided.

## Funding

This project was funded by the SCRI-LIMCR GmbH. The AGMT MBC-registry is supported by grants from Roche, Daiichi Sankyo, Novartis, Pfizer, Caris Life Science, Eli Lilly, Seagen, Gilead, AstraZeneca, and Stemline. Parts of the analysis work were supported by the Deutsche Forschungsgemeinschaft (DFG, German Research Foundation) under Germany’s Excellence Strategy (project IDs 390685813 - EXC 2047 and 390873048 - EXC 2151) and through Metaflammation, project ID 432325352 – SFB 1454, German (project IDs 01EQ2404D and 031L0308B), the European Union via ERC grant INTEGRATE (grant no 101126146) and by the University of Bonn via the Schlegel professorship to the laboratory of J.H..

The supporters did not have any involvement in study design, selection or enrolment of patients, data collection, storage, analysis, interpretation of the data, preparation of the manuscript or the decision to submit the manuscript for publication.

## Author contributions

Concept and design: M.V., G.R., R.G. and J.H.. Acquisition of data: G.R., S.P.G., C.F.S., T.S., F.R., C.H., P.P., S.U., R.B., S.H., D.E., C.A.S., A.F.Z., M.S., J.A., R.P., R.G.. Model development and statistical analysis: M.V., N.B. and J.H.. Interpretation of results: M.V., G.R., S.P.G., R.B., D.E., R.G., J.H.. Acquisition of funding and supervision: R.G. and J.H.. Preparation of first draft: M.V., G.R., and J.H.. All authors critically reviewed and revised the manuscript and approved the final version for submission.

## Data and code availability

The analysis code is available at GitHub (github.com/marc-vaisband/MBC-CTMC).

## A Descriptive details of studied patient population

To explore whether breast cancer progression differed by molecular subtype, we compared clinical characteristics across subtype groups. These overall comparisons showed that subtype was associated with age at first diagnosis of metastatic disease (Kruskal–Wallis *p* = 1.96 × 10^−8^), tumour grade among known cases (*χ*^2^ *p* = 1.09 × 10^−204^), menopausal status among known cases (*χ*^2^ *p* = 5.95×10^−8^), whether patients were diagnosed in M0 stage as represented by DFS availability (*χ*^2^ *p* = 9.32 × 10^−8^), DFS among patients with available DFS (Kruskal–Wallis *p* = 3.02 × 10^−40^), and the number of metastatic sites at first diagnosis of metastatic disease (*χ*^2^ *p* = 4.90 × 10^−3^). Post-hoc analyses supported a broadly more favourable clinical profile for luminal-like disease, including older age at first diagnosis of metastatic disease, lower grade in the luminal A like group, and longer DFS in luminal-like subtypes than in TNBC, with luminal A like, luminal B like, and Luminal HER2+ each showing longer DFS than TNBC (*p*_Bonf_ = 1.78 × 10^−36^, 1.70 × 10^−20^, and 1.03 × 10^−12^, respectively).

Metastatic organ localisation also differed substantially by molecular subtype. For metastases observed at any time during the recorded disease trajectory, significant subtype associations were found for bone (*χ*^2^ *p* = 5.02×10^−25^), lung (*χ*^2^ *p* = 1.02×10^−9^), liver (*χ*^2^ *p* = 8.11×10^−3^), lymph nodes (*χ*^2^ *p* = 2.28 × 10^−9^), brain (*χ*^2^ *p* = 7.15 × 10^−39^), and other localisations (*χ*^2^ *p* = 3.88 × 10^−4^). Bone metastases showed the clearest luminal-like enrichment: all luminal-like subtypes had a higher proportion of patients with bone metastases than TNBC patients when considering metastases at any time (*p*_Bonf_ = 1.67 × 10^−18^, 6.45 × 10^−13^, and 6.43 × 10^−6^, respectively). This pattern was also present at first diagnosis of metastatic disease, where bone involvement again differed by subtype (*χ*^2^ *p* = 1.24 × 10^−12^) and was more frequent in each luminal-like subtype than in TNBC (*p*_Bonf_ = 2.11 × 10^−8^, 3.34 × 10^−7^, and 1.02 × 10^−13^, respectively).

Other metastatic localisations showed distinct subtype-associated patterns. Brain metastases at any time were enriched in HER2+ and TNBC disease relative to luminal A like disease, and also in Luminal HER2+ relative to luminal A like disease (*p*_Bonf_ = 4.56 × 10^−25^, 1.20 × 10^−18^, and 7.51 × 10^−18^, respectively). Liver involvement at first diagnosis of metastatic disease was most characteristic of HER2-positive subtypes, with significant differences between luminal A like and both HER2+ and Luminal HER2+ disease (*p*_Bonf_ = 2.00 × 10^−10^ and 2.81 × 10^−10^, respectively). At first diagnosis of metastatic disease, subtype associations were also observed for lung (*χ*^2^ *p* = 3.81 × 10^−2^), liver (*χ*^2^ *p* = 5.64 × 10^−15^), lymph-node (*χ*^2^ *p* = 4.27 × 10^−3^), and brain involvement (*χ*^2^ *p* = 1.90 × 10^−5^), whereas the aggregated other localisation did not differ by subtype (*χ*^2^ *p* = 6.63 × 10^−1^).

The full results of the analyses can be found in the tables below.

**Table S1:** Omnibus tests.

| Tested quantity | $n$ | $p$ |
| --- | --- | --- |
| Grade at diagnosis (1/2/3, among known) | 2062 | $1.09 \times 10^{-204}$ |
| DFS availability | 2506 | $9.32 \times 10^{-8}$ |
| Menopausal status (pre/post, among known) | 2032 | $5.95 \times 10^{-8}$ |
| Number of metastases at first metastatic diagnosis | 2506 | $4.90 \times 10^{-3}$ |
| Metastasis anytime: Bone | 2506 | $5.02 \times 10^{-25}$ |
| Metastasis anytime: Lungs | 2506 | $1.02 \times 10^{-9}$ |
| Metastasis anytime: Liver | 2506 | $8.11 \times 10^{-3}$ |
| Metastasis anytime: Lymph nodes | 2506 | $2.28 \times 10^{-9}$ |
| Metastasis anytime: Brain | 2506 | $7.15 \times 10^{-39}$ |
| Metastasis anytime: Other | 2506 | $3.88 \times 10^{-4}$ |
| Bone at first metastatic diagnosis | 2506 | $1.24 \times 10^{-12}$ |
| Lungs at first metastatic diagnosis | 2506 | $3.81 \times 10^{-2}$ |
| Liver at first metastatic diagnosis | 2506 | $5.64 \times 10^{-15}$ |
| Lymph nodes at first metastatic diagnosis | 2506 | $4.27 \times 10^{-3}$ |
| Brain at first metastatic diagnosis | 2506 | $1.90 \times 10^{-5}$ |
| Other at first metastatic diagnosis | 2506 | $6.63 \times 10^{-1}$ |
| Age at first metastatic diagnosis (among available) | 2506 | $1.96 \times 10^{-8}$ |
| DFS (among available) | 1412 | $3.02 \times 10^{-40}$ |

**Table S2:** Age comparisons.

| Subtype A | Subtype B | $n_A$ | $n_B$ | median<sub>A</sub> | median<sub>B</sub> | $p$ | $P_{\text{Bonf}}$ |
| --- | --- | --- | --- | --- | --- | --- | --- |
| Luminal A like | TNBC | 916 | 380 | 65.0 | 60.0 | $9.69 \times 10^{-9}$ | $9.69 \times 10^{-8}$ |
| Luminal A like | HER2+ | 916 | 174 | 65.0 | 59.5 | $4.70 \times 10^{-5}$ | $4.70 \times 10^{-4}$ |
| Luminal A like | Luminal B like | 916 | 730 | 65.0 | 63.0 | $1.31 \times 10^{-3}$ | $1.31 \times 10^{-2}$ |
| Luminal A like | Luminal HER2+ | 916 | 306 | 65.0 | 63.0 | $1.90 \times 10^{-3}$ | $1.90 \times 10^{-2}$ |
| Luminal B like | TNBC | 730 | 380 | 63.0 | 60.0 | $5.33 \times 10^{-3}$ | $5.33 \times 10^{-2}$ |
| Luminal B like | HER2+ | 730 | 174 | 63.0 | 59.5 | $5.65 \times 10^{-2}$ | $5.65 \times 10^{-1}$ |
| HER2+ | TNBC | 174 | 380 | 59.5 | 60.0 | $9.91 \times 10^{-1}$ | $1.00 \times 10^0$ |
| Luminal B like | Luminal HER2+ | 730 | 306 | 63.0 | 63.0 | $4.30 \times 10^{-1}$ | $1.00 \times 10^0$ |
| Luminal HER2+ | HER2+ | 306 | 174 | 63.0 | 59.5 | $2.47 \times 10^{-1}$ | $1.00 \times 10^0$ |
| Luminal HER2+ | TNBC | 306 | 380 | 63.0 | 60.0 | $1.10 \times 10^{-1}$ | $1.00 \times 10^0$ |

**Table S3:** Menopausal status comparisons.

| Subtype A | Subtype B | $n_A$ | $n_B$ | (Pre-menopausal, Post-menopausal)<sub>A</sub> | (Pre-menopausal, Post-menopausal)<sub>B</sub> | $p$ | $P_{\text{Bonf}}$ |
| --- | --- | --- | --- | --- | --- | --- | --- |
| Luminal A like | HER2+ | 748 | 133 | 10.0%, 90.0% | 25.6%, 74.4% | $1.11 \times 10^{-6}$ | $1.11 \times 10^{-5}$ |
| Luminal A like | TNBC | 748 | 296 | 10.0%, 90.0% | 21.3%, 78.7% | $2.15 \times 10^{-6}$ | $2.15 \times 10^{-5}$ |
| Luminal A like | Luminal B like | 748 | 613 | 10.0%, 90.0% | 17.9%, 82.1% | $3.17 \times 10^{-5}$ | $3.17 \times 10^{-4}$ |
| Luminal A like | Luminal HER2+ | 748 | 242 | 10.0%, 90.0% | 20.2%, 79.8% | $4.83 \times 10^{-5}$ | $4.83 \times 10^{-4}$ |
| Luminal B like | HER2+ | 613 | 133 | 17.9%, 82.1% | 25.6%, 74.4% | $5.78 \times 10^{-2}$ | $5.78 \times 10^{-1}$ |
| HER2+ | TNBC | 133 | 296 | 25.6%, 74.4% | 21.3%, 78.7% | $3.92 \times 10^{-1}$ | $1.00 \times 10^0$ |
| Luminal B like | Luminal HER2+ | 613 | 242 | 17.9%, 82.1% | 20.2%, 79.8% | $4.95 \times 10^{-1}$ | $1.00 \times 10^0$ |
| Luminal B like | TNBC | 613 | 296 | 17.9%, 82.1% | 21.3%, 78.7% | $2.66 \times 10^{-1}$ | $1.00 \times 10^0$ |
| Luminal HER2+ | HER2+ | 242 | 133 | 20.2%, 79.8% | 25.6%, 74.4% | $2.91 \times 10^{-1}$ | $1.00 \times 10^0$ |
| Luminal HER2+ | TNBC | 242 | 296 | 20.2%, 79.8% | 21.3%, 78.7% | $8.51 \times 10^{-1}$ | $1.00 \times 10^0$ |

**Table S4:**
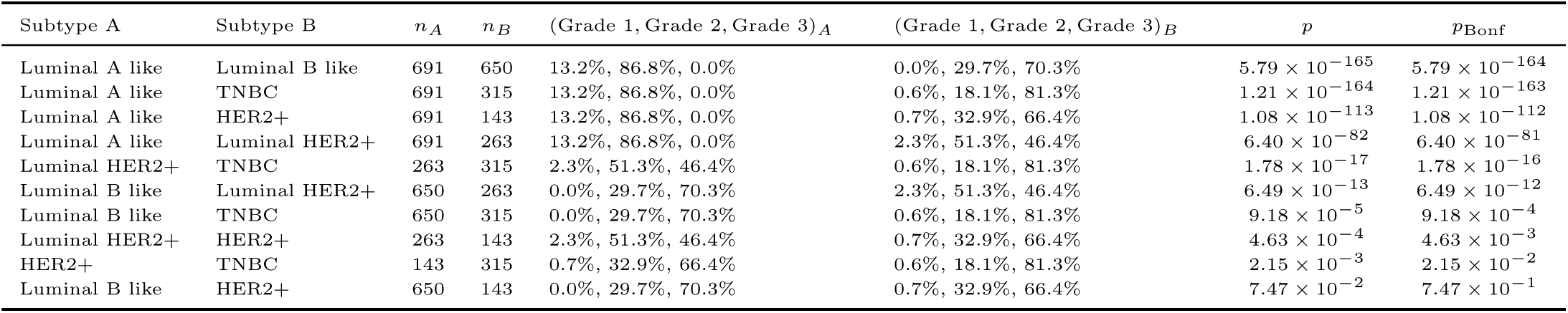
Grade comparisons.

**Table S5:** Bone metastasis anytime comparisons.

| Subtype A | Subtype B | $n_A$ | $n_B$ | (Yes, No)$_A$ | (Yes, No)$_B$ | $p$ | $P_{\text{Bonf}}$ |
| --- | --- | --- | --- | --- | --- | --- | --- |
| Luminal A like | TNBC | 916 | 380 | 77.9%, 22.1% | 52.6%, 47.4% | $1.67 \times 10^{-19}$ | $1.67 \times 10^{-18}$ |
| Luminal B like | TNBC | 730 | 380 | 75.1%, 24.9% | 52.6%, 47.4% | $6.45 \times 10^{-14}$ | $6.45 \times 10^{-13}$ |
| Luminal A like | HER2+ | 916 | 174 | 77.9%, 22.1% | 51.7%, 48.3% | $1.13 \times 10^{-12}$ | $1.13 \times 10^{-11}$ |
| Luminal B like | HER2+ | 730 | 174 | 75.1%, 24.9% | 51.7%, 48.3% | $2.24 \times 10^{-9}$ | $2.24 \times 10^{-8}$ |
| Luminal HER2+ | TNBC | 306 | 380 | 71.6%, 28.4% | 52.6%, 47.4% | $6.43 \times 10^{-7}$ | $6.43 \times 10^{-6}$ |
| Luminal HER2+ | HER2+ | 306 | 174 | 71.6%, 28.4% | 51.7%, 48.3% | $2.00 \times 10^{-5}$ | $2.00 \times 10^{-4}$ |
| Luminal A like | Luminal HER2+ | 916 | 306 | 77.9%, 22.1% | 71.6%, 28.4% | $2.81 \times 10^{-2}$ | $2.81 \times 10^{-1}$ |
| HER2+ | TNBC | 174 | 380 | 51.7%, 48.3% | 52.6%, 47.4% | $9.15 \times 10^{-1}$ | $1.00 \times 10^0$ |
| Luminal A like | Luminal B like | 916 | 730 | 77.9%, 22.1% | 75.1%, 24.9% | $1.89 \times 10^{-1}$ | $1.00 \times 10^0$ |
| Luminal B like | Luminal HER2+ | 730 | 306 | 75.1%, 24.9% | 71.6%, 28.4% | $2.74 \times 10^{-1}$ | $1.00 \times 10^0$ |

**Table S6:** Bone metastasis at first diagnosis of metastatic disease comparisons.

| Subtype A | Subtype B | $n_A$ | $n_B$ | (Yes, No)$_A$ | (Yes, No)$_B$ | $p$ | $P_{\text{Bonf}}$ |
| --- | --- | --- | --- | --- | --- | --- | --- |
| Luminal HER2+ | TNBC | 306 | 380 | 40.2%, 59.8% | 13.9%, 86.1% | $1.02 \times 10^{-14}$ | $1.02 \times 10^{-13}$ |
| Luminal A like | TNBC | 916 | 380 | 30.0%, 70.0% | 13.9%, 86.1% | $2.11 \times 10^{-9}$ | $2.11 \times 10^{-8}$ |
| Luminal B like | TNBC | 730 | 380 | 29.0%, 71.0% | 13.9%, 86.1% | $3.34 \times 10^{-8}$ | $3.34 \times 10^{-7}$ |
| HER2+ | TNBC | 174 | 380 | 26.4%, 73.6% | 13.9%, 86.1% | $5.77 \times 10^{-4}$ | $5.77 \times 10^{-3}$ |
| Luminal B like | Luminal HER2+ | 730 | 306 | 29.0%, 71.0% | 40.2%, 59.8% | $6.06 \times 10^{-4}$ | $6.06 \times 10^{-3}$ |
| Luminal A like | Luminal HER2+ | 916 | 306 | 30.0%, 70.0% | 40.2%, 59.8% | $1.29 \times 10^{-3}$ | $1.29 \times 10^{-2}$ |
| Luminal HER2+ | HER2+ | 306 | 174 | 40.2%, 59.8% | 26.4%, 73.6% | $3.34 \times 10^{-3}$ | $3.34 \times 10^{-2}$ |
| Luminal A like | HER2+ | 916 | 174 | 30.0%, 70.0% | 26.4%, 73.6% | $3.90 \times 10^{-1}$ | $1.00 \times 10^0$ |
| Luminal A like | Luminal B like | 916 | 730 | 30.0%, 70.0% | 29.0%, 71.0% | $7.05 \times 10^{-1}$ | $1.00 \times 10^0$ |
| Luminal B like | HER2+ | 730 | 174 | 29.0%, 71.0% | 26.4%, 73.6% | $5.55 \times 10^{-1}$ | $1.00 \times 10^0$ |

**Table S7:** Lungs metastasis anytime comparisons.

| Subtype A | Subtype B | $n_A$ | $n_B$ | (Yes, No)$_A$ | (Yes, No)$_B$ | $p$ | $P_{\text{Bonf}}$ |
| --- | --- | --- | --- | --- | --- | --- | --- |
| Luminal A like | TNBC | 916 | 380 | 32.2%, 67.8% | 51.6%, 48.4% | $9.04 \times 10^{-11}$ | $9.04 \times 10^{-10}$ |
| Luminal A like | Luminal B like | 916 | 730 | 32.2%, 67.8% | 41.8%, 58.2% | $7.54 \times 10^{-5}$ | $7.54 \times 10^{-4}$ |
| Luminal A like | Luminal HER2+ | 916 | 306 | 32.2%, 67.8% | 42.5%, 57.5% | $1.38 \times 10^{-3}$ | $1.38 \times 10^{-2}$ |
| Luminal A like | HER2+ | 916 | 174 | 32.2%, 67.8% | 44.8%, 55.2% | $1.75 \times 10^{-3}$ | $1.75 \times 10^{-2}$ |
| Luminal B like | TNBC | 730 | 380 | 41.8%, 58.2% | 51.6%, 48.4% | $2.30 \times 10^{-3}$ | $2.30 \times 10^{-2}$ |
| Luminal HER2+ | TNBC | 306 | 380 | 42.5%, 57.5% | 51.6%, 48.4% | $2.18 \times 10^{-2}$ | $2.18 \times 10^{-1}$ |
| HER2+ | TNBC | 174 | 380 | 44.8%, 55.2% | 51.6%, 48.4% | $1.66 \times 10^{-1}$ | $1.00 \times 10^0$ |
| Luminal B like | HER2+ | 730 | 174 | 41.8%, 58.2% | 44.8%, 55.2% | $5.19 \times 10^{-1}$ | $1.00 \times 10^0$ |
| Luminal B like | Luminal HER2+ | 730 | 306 | 41.8%, 58.2% | 42.5%, 57.5% | $8.89 \times 10^{-1}$ | $1.00 \times 10^0$ |
| Luminal HER2+ | HER2+ | 306 | 174 | 42.5%, 57.5% | 44.8%, 55.2% | $6.87 \times 10^{-1}$ | $1.00 \times 10^0$ |

**Table S8:** Lungs metastasis at first diagnosis of metastatic disease comparisons.

| Subtype A | Subtype B | $n_A$ | $n_B$ | (Yes, No)$_A$ | (Yes, No)$_B$ | $p$ | $P_{\text{Bonf}}$ |
| --- | --- | --- | --- | --- | --- | --- | --- |
| Luminal A like | Luminal HER2+ | 916 | 306 | 10.3%, 89.7% | 15.7%, 84.3% | $1.39 \times 10^{-2}$ | $1.39 \times 10^{-1}$ |
| Luminal A like | HER2+ | 916 | 174 | 10.3%, 89.7% | 16.7%, 83.3% | $2.05 \times 10^{-2}$ | $2.05 \times 10^{-1}$ |
| HER2+ | TNBC | 174 | 380 | 16.7%, 83.3% | 11.8%, 88.2% | $1.57 \times 10^{-1}$ | $1.00 \times 10^0$ |
| Luminal A like | Luminal B like | 916 | 730 | 10.3%, 89.7% | 11.8%, 88.2% | $3.67 \times 10^{-1}$ | $1.00 \times 10^0$ |
| Luminal A like | TNBC | 916 | 380 | 10.3%, 89.7% | 11.8%, 88.2% | $4.60 \times 10^{-1}$ | $1.00 \times 10^0$ |
| Luminal B like | HER2+ | 730 | 174 | 11.8%, 88.2% | 16.7%, 83.3% | $1.07 \times 10^{-1}$ | $1.00 \times 10^0$ |
| Luminal B like | Luminal HER2+ | 730 | 306 | 11.8%, 88.2% | 15.7%, 84.3% | $1.08 \times 10^{-1}$ | $1.00 \times 10^0$ |
| Luminal B like | TNBC | 730 | 380 | 11.8%, 88.2% | 11.8%, 88.2% | $1.00 \times 10^0$ | $1.00 \times 10^0$ |
| Luminal HER2+ | HER2+ | 306 | 174 | 15.7%, 84.3% | 16.7%, 83.3% | $8.79 \times 10^{-1}$ | $1.00 \times 10^0$ |
| Luminal HER2+ | TNBC | 306 | 380 | 15.7%, 84.3% | 11.8%, 88.2% | $1.77 \times 10^{-1}$ | $1.00 \times 10^0$ |

**Table S9:** Liver metastasis anytime comparisons.

| Subtype A | Subtype B | $n_A$ | $n_B$ | (Yes, No)$_A$ | (Yes, No)$_B$ | $p$ | $P_{\text{Bonf}}$ |
| --- | --- | --- | --- | --- | --- | --- | --- |
| Luminal A like | Luminal B like | 916 | 730 | 44.7%, 55.3% | 53.3%, 46.7% | $5.96 \times 10^{-4}$ | $5.96 \times 10^{-3}$ |
| Luminal B like | TNBC | 730 | 380 | 53.3%, 46.7% | 45.8%, 54.2% | $2.10 \times 10^{-2}$ | $2.10 \times 10^{-1}$ |
| Luminal B like | Luminal HER2+ | 730 | 306 | 53.3%, 46.7% | 45.4%, 54.6% | $2.50 \times 10^{-2}$ | $2.50 \times 10^{-1}$ |
| HER2+ | TNBC | 174 | 380 | 47.1%, 52.9% | 45.8%, 54.2% | $8.41 \times 10^{-1}$ | $1.00 \times 10^0$ |
| Luminal A like | HER2+ | 916 | 174 | 44.7%, 55.3% | 47.1%, 52.9% | $6.04 \times 10^{-1}$ | $1.00 \times 10^0$ |
| Luminal A like | Luminal HER2+ | 916 | 306 | 44.7%, 55.3% | 45.4%, 54.6% | $8.66 \times 10^{-1}$ | $1.00 \times 10^0$ |
| Luminal A like | TNBC | 916 | 380 | 44.7%, 55.3% | 45.8%, 54.2% | $7.54 \times 10^{-1}$ | $1.00 \times 10^0$ |
| Luminal B like | HER2+ | 730 | 174 | 53.3%, 46.7% | 47.1%, 52.9% | $1.68 \times 10^{-1}$ | $1.00 \times 10^0$ |
| Luminal HER2+ | HER2+ | 306 | 174 | 45.4%, 54.6% | 47.1%, 52.9% | $7.92 \times 10^{-1}$ | $1.00 \times 10^0$ |
| Luminal HER2+ | TNBC | 306 | 380 | 45.4%, 54.6% | 45.8%, 54.2% | $9.85 \times 10^{-1}$ | $1.00 \times 10^0$ |

**Table S10:** Liver metastasis at first diagnosis of metastatic disease comparisons.

| Subtype A | Subtype B | $n_A$ | $n_B$ | (Yes, No)$_A$ | (Yes, No)$_B$ | $p$ | $P_{\text{Bonf}}$ |
| --- | --- | --- | --- | --- | --- | --- | --- |
| Luminal A like | HER2+ | 916 | 174 | 6.0%, 94.0% | 21.8%, 78.2% | $2.00 \times 10^{-11}$ | $2.00 \times 10^{-10}$ |
| Luminal A like | Luminal HER2+ | 916 | 306 | 6.0%, 94.0% | 19.0%, 81.0% | $2.81 \times 10^{-11}$ | $2.81 \times 10^{-10}$ |
| HER2+ | TNBC | 174 | 380 | 21.8%, 78.2% | 7.1%, 92.9% | $1.18 \times 10^{-6}$ | $1.18 \times 10^{-5}$ |
| Luminal HER2+ | TNBC | 306 | 380 | 19.0%, 81.0% | 7.1%, 92.9% | $4.98 \times 10^{-6}$ | $4.98 \times 10^{-5}$ |
| Luminal B like | HER2+ | 730 | 174 | 10.8%, 89.2% | 21.8%, 78.2% | $1.67 \times 10^{-4}$ | $1.67 \times 10^{-3}$ |
| Luminal A like | Luminal B like | 916 | 730 | 6.0%, 94.0% | 10.8%, 89.2% | $5.40 \times 10^{-4}$ | $5.40 \times 10^{-3}$ |
| Luminal B like | Luminal HER2+ | 730 | 306 | 10.8%, 89.2% | 19.0%, 81.0% | $6.16 \times 10^{-4}$ | $6.16 \times 10^{-3}$ |
| Luminal B like | TNBC | 730 | 380 | 10.8%, 89.2% | 7.1%, 92.9% | $5.86 \times 10^{-2}$ | $5.86 \times 10^{-1}$ |
| Luminal A like | TNBC | 916 | 380 | 6.0%, 94.0% | 7.1%, 92.9% | $5.38 \times 10^{-1}$ | $1.00 \times 10^0$ |
| Luminal HER2+ | HER2+ | 306 | 174 | 19.0%, 81.0% | 21.8%, 78.2% | $5.22 \times 10^{-1}$ | $1.00 \times 10^0$ |

**Table S11:** Lymph nodes metastasis anytime comparisons.

| Subtype A | Subtype B | $n_A$ | $n_B$ | (Yes, No)$_A$ | (Yes, No)$_B$ | $p$ | $P_{\text{Bonf}}$ |
| --- | --- | --- | --- | --- | --- | --- | --- |
| Luminal A like | TNBC | 916 | 380 | 36.5%, 63.5% | 56.1%, 43.9% | $1.21 \times 10^{-10}$ | $1.21 \times 10^{-9}$ |
| Luminal A like | Luminal B like | 916 | 730 | 36.5%, 63.5% | 46.7%, 53.3% | $3.33 \times 10^{-5}$ | $3.33 \times 10^{-4}$ |
| Luminal HER2+ | TNBC | 306 | 380 | 43.8%, 56.2% | 56.1%, 43.9% | $1.83 \times 10^{-3}$ | $1.83 \times 10^{-2}$ |
| Luminal B like | TNBC | 730 | 380 | 46.7%, 53.3% | 56.1%, 43.9% | $3.85 \times 10^{-3}$ | $3.85 \times 10^{-2}$ |
| Luminal A like | HER2+ | 916 | 174 | 36.5%, 63.5% | 46.6%, 53.4% | $1.52 \times 10^{-2}$ | $1.52 \times 10^{-1}$ |
| Luminal A like | Luminal HER2+ | 916 | 306 | 36.5%, 63.5% | 43.8%, 56.2% | $2.67 \times 10^{-2}$ | $2.67 \times 10^{-1}$ |
| HER2+ | TNBC | 174 | 380 | 46.6%, 53.4% | 56.1%, 43.9% | $4.68 \times 10^{-2}$ | $4.68 \times 10^{-1}$ |
| Luminal B like | HER2+ | 730 | 174 | 46.7%, 53.3% | 46.6%, 53.4% | $1.00 \times 10^0$ | $1.00 \times 10^0$ |
| Luminal B like | Luminal HER2+ | 730 | 306 | 46.7%, 53.3% | 43.8%, 56.2% | $4.28 \times 10^{-1}$ | $1.00 \times 10^0$ |
| Luminal HER2+ | HER2+ | 306 | 174 | 43.8%, 56.2% | 46.6%, 53.4% | $6.25 \times 10^{-1}$ | $1.00 \times 10^0$ |

**Table S12:**
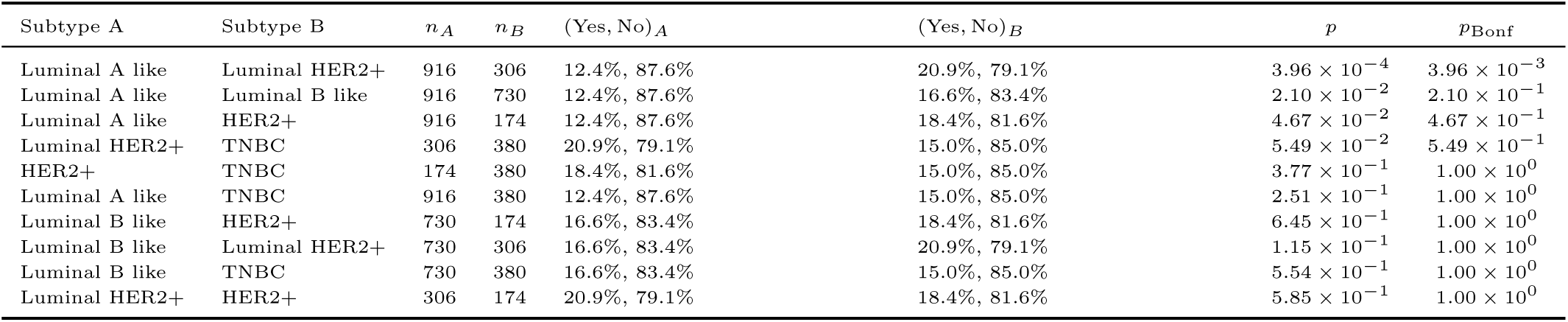
Lymph nodes metastasis at first diagnosis of metastatic disease comparisons.

**Table S13:** Brain metastasis anytime comparisons.

| Subtype A | Subtype B | $n_A$ | $n_B$ | (Yes, No)$_A$ | (Yes, No)$_B$ | $p$ | $P_{\text{Bonf}}$ |
| --- | --- | --- | --- | --- | --- | --- | --- |
| Luminal A like | HER2+ | 916 | 174 | 9.1%, 90.9% | 39.7%, 60.3% | $4.56 \times 10^{-26}$ | $4.56 \times 10^{-25}$ |
| Luminal A like | TNBC | 916 | 380 | 9.1%, 90.9% | 28.9%, 71.1% | $1.20 \times 10^{-19}$ | $1.20 \times 10^{-18}$ |
| Luminal A like | Luminal HER2+ | 916 | 306 | 9.1%, 90.9% | 29.7%, 70.3% | $7.51 \times 10^{-19}$ | $7.51 \times 10^{-18}$ |
| Luminal B like | HER2+ | 730 | 174 | 12.1%, 87.9% | 39.7%, 60.3% | $1.53 \times 10^{-17}$ | $1.53 \times 10^{-16}$ |
| Luminal B like | TNBC | 730 | 380 | 12.1%, 87.9% | 28.9%, 71.1% | $5.46 \times 10^{-12}$ | $5.46 \times 10^{-11}$ |
| Luminal B like | Luminal HER2+ | 730 | 306 | 12.1%, 87.9% | 29.7%, 70.3% | $1.22 \times 10^{-11}$ | $1.22 \times 10^{-10}$ |
| HER2+ | TNBC | 174 | 380 | 39.7%, 60.3% | 28.9%, 71.1% | $1.62 \times 10^{-2}$ | $1.62 \times 10^{-1}$ |
| Luminal HER2+ | HER2+ | 306 | 174 | 29.7%, 70.3% | 39.7%, 60.3% | $3.44 \times 10^{-2}$ | $3.44 \times 10^{-1}$ |
| Luminal A like | Luminal B like | 916 | 730 | 9.1%, 90.9% | 12.1%, 87.9% | $5.79 \times 10^{-2}$ | $5.79 \times 10^{-1}$ |
| Luminal HER2+ | TNBC | 306 | 380 | 29.7%, 70.3% | 28.9%, 71.1% | $8.87 \times 10^{-1}$ | $1.00 \times 10^0$ |

**Table S14:** Brain metastasis at first diagnosis of metastatic disease comparisons.

| Subtype A | Subtype B | $n_A$ | $n_B$ | (Yes, No)$_A$ | (Yes, No)$_B$ | $p$ | $P_{\text{Bonf}}$ |
| --- | --- | --- | --- | --- | --- | --- | --- |
| Luminal A like | Luminal HER2+ | 916 | 306 | 0.5%, 99.5% | 3.9%, 96.1% | $8.99 \times 10^{-5}$ | $8.99 \times 10^{-4}$ |
| Luminal B like | Luminal HER2+ | 730 | 306 | 0.8%, 99.2% | 3.9%, 96.1% | $1.27 \times 10^{-3}$ | $1.27 \times 10^{-2}$ |
| Luminal HER2+ | TNBC | 306 | 380 | 3.9%, 96.1% | 0.8%, 99.2% | $1.15 \times 10^{-2}$ | $1.15 \times 10^{-1}$ |
| Luminal A like | HER2+ | 916 | 174 | 0.5%, 99.5% | 2.9%, 97.1% | $1.25 \times 10^{-2}$ | $1.25 \times 10^{-1}$ |
| Luminal B like | HER2+ | 730 | 174 | 0.8%, 99.2% | 2.9%, 97.1% | $4.24 \times 10^{-2}$ | $4.24 \times 10^{-1}$ |
| HER2+ | TNBC | 174 | 380 | 2.9%, 97.1% | 0.8%, 99.2% | $1.16 \times 10^{-1}$ | $1.00 \times 10^0$ |
| Luminal A like | Luminal B like | 916 | 730 | 0.5%, 99.5% | 0.8%, 99.2% | $5.52 \times 10^{-1}$ | $1.00 \times 10^0$ |
| Luminal A like | TNBC | 916 | 380 | 0.5%, 99.5% | 0.8%, 99.2% | $6.99 \times 10^{-1}$ | $1.00 \times 10^0$ |
| Luminal B like | TNBC | 730 | 380 | 0.8%, 99.2% | 0.8%, 99.2% | $1.00 \times 10^0$ | $1.00 \times 10^0$ |
| Luminal HER2+ | HER2+ | 306 | 174 | 3.9%, 96.1% | 2.9%, 97.1% | $7.34 \times 10^{-1}$ | $1.00 \times 10^0$ |

**Table S15:** Other metastasis anytime comparisons.

| Subtype A | Subtype B | $n_A$ | $n_B$ | (Yes, No)$_A$ | (Yes, No)$_B$ | $p$ | $P_{\text{Bonf}}$ |
| --- | --- | --- | --- | --- | --- | --- | --- |
| HER2+ | TNBC | 174 | 380 | 37.9%, 62.1% | 55.3%, 44.7% | $2.19 \times 10^{-4}$ | $2.19 \times 10^{-3}$ |
| Luminal HER2+ | TNBC | 306 | 380 | 42.5%, 57.5% | 55.3%, 44.7% | $1.15 \times 10^{-3}$ | $1.15 \times 10^{-2}$ |
| Luminal B like | HER2+ | 730 | 174 | 50.5%, 49.5% | 37.9%, 62.1% | $3.63 \times 10^{-3}$ | $3.63 \times 10^{-2}$ |
| Luminal A like | HER2+ | 916 | 174 | 49.7%, 50.3% | 37.9%, 62.1% | $5.79 \times 10^{-3}$ | $5.79 \times 10^{-2}$ |
| Luminal B like | Luminal HER2+ | 730 | 306 | 50.5%, 49.5% | 42.5%, 57.5% | $2.13 \times 10^{-2}$ | $2.13 \times 10^{-1}$ |
| Luminal A like | Luminal HER2+ | 916 | 306 | 49.7%, 50.3% | 42.5%, 57.5% | $3.46 \times 10^{-2}$ | $3.46 \times 10^{-1}$ |
| Luminal A like | TNBC | 916 | 380 | 49.7%, 50.3% | 55.3%, 44.7% | $7.64 \times 10^{-2}$ | $7.64 \times 10^{-1}$ |
| Luminal A like | Luminal B like | 916 | 730 | 49.7%, 50.3% | 50.5%, 49.5% | $7.62 \times 10^{-1}$ | $1.00 \times 10^0$ |
| Luminal B like | TNBC | 730 | 380 | 50.5%, 49.5% | 55.3%, 44.7% | $1.53 \times 10^{-1}$ | $1.00 \times 10^0$ |
| Luminal HER2+ | HER2+ | 306 | 174 | 42.5%, 57.5% | 37.9%, 62.1% | $3.79 \times 10^{-1}$ | $1.00 \times 10^0$ |

**Table S16:**
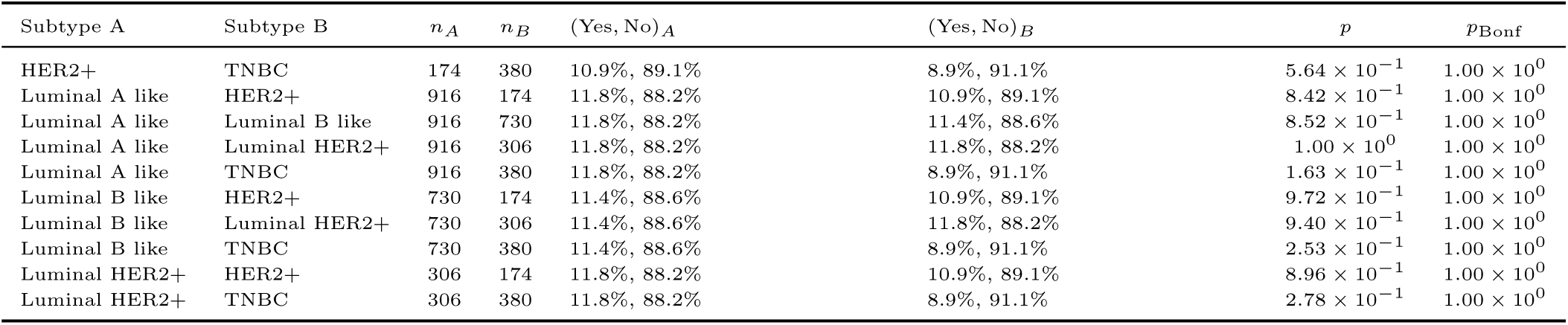
Other metastasis at first diagnosis of metastatic disease comparisons.

| Subtype A | Subtype B | $n_A$ | $n_B$ | (Yes, No)$_A$ | (Yes, No)$_B$ | $p$ | $p_{\text{Bonf}}$ |
| --- | --- | --- | --- | --- | --- | --- | --- |
| HER2+ | TNBC | 174 | 380 | 10.9%, 89.1% | 8.9%, 91.1% | $5.64 \times 10^{-1}$ | $1.00 \times 10^0$ |
| Luminal A like | HER2+ | 916 | 174 | 11.8%, 88.2% | 10.9%, 89.1% | $8.42 \times 10^{-1}$ | $1.00 \times 10^0$ |
| Luminal A like | Luminal B like | 916 | 730 | 11.8%, 88.2% | 11.4%, 88.6% | $8.52 \times 10^{-1}$ | $1.00 \times 10^0$ |
| Luminal A like | Luminal HER2+ | 916 | 306 | 11.8%, 88.2% | 11.8%, 88.2% | $1.00 \times 10^0$ | $1.00 \times 10^0$ |
| Luminal A like | TNBC | 916 | 380 | 11.8%, 88.2% | 8.9%, 91.1% | $1.63 \times 10^{-1}$ | $1.00 \times 10^0$ |
| Luminal B like | HER2+ | 730 | 174 | 11.4%, 88.6% | 10.9%, 89.1% | $9.72 \times 10^{-1}$ | $1.00 \times 10^0$ |
| Luminal B like | Luminal HER2+ | 730 | 306 | 11.4%, 88.6% | 11.8%, 88.2% | $9.40 \times 10^{-1}$ | $1.00 \times 10^0$ |
| Luminal B like | TNBC | 730 | 380 | 11.4%, 88.6% | 8.9%, 91.1% | $2.53 \times 10^{-1}$ | $1.00 \times 10^0$ |
| Luminal HER2+ | HER2+ | 306 | 174 | 11.8%, 88.2% | 10.9%, 89.1% | $8.96 \times 10^{-1}$ | $1.00 \times 10^0$ |
| Luminal HER2+ | TNBC | 306 | 380 | 11.8%, 88.2% | 8.9%, 91.1% | $2.78 \times 10^{-1}$ | $1.00 \times 10^0$ |

**Table S17:**
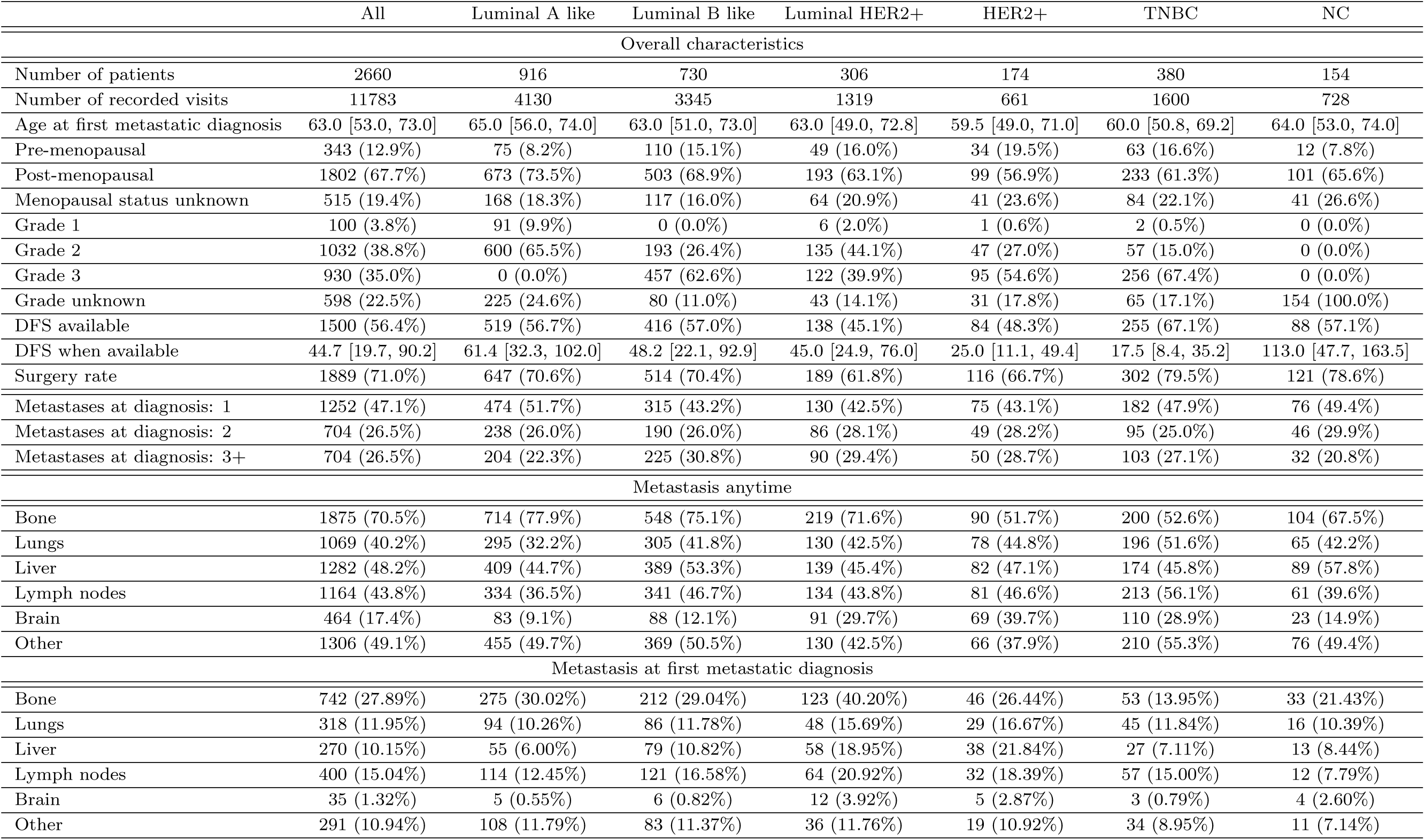
Characteristics of the studied patient population. Categorical variables are given as counts (relative proportions). Continuous variables are reported as median [interquartile range].

|  | All | Luminal A like | Luminal B like | Luminal HER2+ | HER2+ | TNBC | NC |
| --- | --- | --- | --- | --- | --- | --- | --- |
| Overall characteristics |  |  |  |  |  |  |  |
| Number of patients | 2660 | 916 | 730 | 306 | 174 | 380 | 154 |
| Number of recorded visits | 11783 | 4130 | 3345 | 1319 | 661 | 1600 | 728 |
| Age at first metastatic diagnosis | 63.0 [53.0, 73.0] | 65.0 [56.0, 74.0] | 63.0 [51.0, 73.0] | 63.0 [49.0, 72.8] | 59.5 [49.0, 71.0] | 60.0 [50.8, 69.2] | 64.0 [53.0, 74.0] |
| Pre-menopausal | 343 (12.9%) | 75 (8.2%) | 110 (15.1%) | 49 (16.0%) | 34 (19.5%) | 63 (16.6%) | 12 (7.8%) |
| Post-menopausal | 1802 (67.7%) | 673 (73.5%) | 503 (68.9%) | 193 (63.1%) | 99 (56.9%) | 233 (61.3%) | 101 (65.6%) |
| Menopausal status unknown | 515 (19.4%) | 168 (18.3%) | 117 (16.0%) | 64 (20.9%) | 41 (23.6%) | 84 (22.1%) | 41 (26.6%) |
| Grade 1 | 100 (3.8%) | 91 (9.9%) | 0 (0.0%) | 6 (2.0%) | 1 (0.6%) | 2 (0.5%) | 0 (0.0%) |
| Grade 2 | 1032 (38.8%) | 600 (65.5%) | 193 (26.4%) | 135 (44.1%) | 47 (27.0%) | 57 (15.0%) | 0 (0.0%) |
| Grade 3 | 930 (35.0%) | 0 (0.0%) | 457 (62.6%) | 122 (39.9%) | 95 (54.6%) | 256 (67.4%) | 0 (0.0%) |
| Grade unknown | 598 (22.5%) | 225 (24.6%) | 80 (11.0%) | 43 (14.1%) | 31 (17.8%) | 65 (17.1%) | 154 (100.0%) |
| DFS available | 1500 (56.4%) | 519 (56.7%) | 416 (57.0%) | 138 (45.1%) | 84 (48.3%) | 255 (67.1%) | 88 (57.1%) |
| DFS when available | 44.7 [19.7, 90.2] | 61.4 [32.3, 102.0] | 48.2 [22.1, 92.9] | 45.0 [24.9, 76.0] | 25.0 [11.1, 49.4] | 17.5 [8.4, 35.2] | 113.0 [47.7, 163.5] |
| Surgery rate | 1889 (71.0%) | 647 (70.6%) | 514 (70.4%) | 189 (61.8%) | 116 (66.7%) | 302 (79.5%) | 121 (78.6%) |
| Metastases at diagnosis: 1 | 1252 (47.1%) | 474 (51.7%) | 315 (43.2%) | 130 (42.5%) | 75 (43.1%) | 182 (47.9%) | 76 (49.4%) |
| Metastases at diagnosis: 2 | 704 (26.5%) | 238 (26.0%) | 190 (26.0%) | 86 (28.1%) | 49 (28.2%) | 95 (25.0%) | 46 (29.9%) |
| Metastases at diagnosis: 3+ | 704 (26.5%) | 204 (22.3%) | 225 (30.8%) | 90 (29.4%) | 50 (28.7%) | 103 (27.1%) | 32 (20.8%) |
| Metastasis anytime |  |  |  |  |  |  |  |
| Bone | 1875 (70.5%) | 714 (77.9%) | 548 (75.1%) | 219 (71.6%) | 90 (51.7%) | 200 (52.6%) | 104 (67.5%) |
| Lungs | 1069 (40.2%) | 295 (32.2%) | 305 (41.8%) | 130 (42.5%) | 78 (44.8%) | 196 (51.6%) | 65 (42.2%) |
| Liver | 1282 (48.2%) | 409 (44.7%) | 389 (53.3%) | 139 (45.4%) | 82 (47.1%) | 174 (45.8%) | 89 (57.8%) |
| Lymph nodes | 1164 (43.8%) | 334 (36.5%) | 341 (46.7%) | 134 (43.8%) | 81 (46.6%) | 213 (56.1%) | 61 (39.6%) |
| Brain | 464 (17.4%) | 83 (9.1%) | 88 (12.1%) | 91 (29.7%) | 69 (39.7%) | 110 (28.9%) | 23 (14.9%) |
| Other | 1306 (49.1%) | 455 (49.7%) | 369 (50.5%) | 130 (42.5%) | 66 (37.9%) | 210 (55.3%) | 76 (49.4%) |
| Metastasis at first metastatic diagnosis |  |  |  |  |  |  |  |
| Bone | 742 (27.89%) | 275 (30.02%) | 212 (29.04%) | 123 (40.20%) | 46 (26.44%) | 53 (13.95%) | 33 (21.43%) |
| Lungs | 318 (11.95%) | 94 (10.26%) | 86 (11.78%) | 48 (15.69%) | 29 (16.67%) | 45 (11.84%) | 16 (10.39%) |
| Liver | 270 (10.15%) | 55 (6.00%) | 79 (10.82%) | 58 (18.95%) | 38 (21.84%) | 27 (7.11%) | 13 (8.44%) |
| Lymph nodes | 400 (15.04%) | 114 (12.45%) | 121 (16.58%) | 64 (20.92%) | 32 (18.39%) | 57 (15.00%) | 12 (7.79%) |
| Brain | 35 (1.32%) | 5 (0.55%) | 6 (0.82%) | 12 (3.92%) | 5 (2.87%) | 3 (0.79%) | 4 (2.60%) |
| Other | 291 (10.94%) | 108 (11.79%) | 83 (11.37%) | 36 (11.76%) | 19 (10.92%) | 34 (8.95%) | 11 (7.14%) |

### A.1 Therapies received

**Figure S1:**
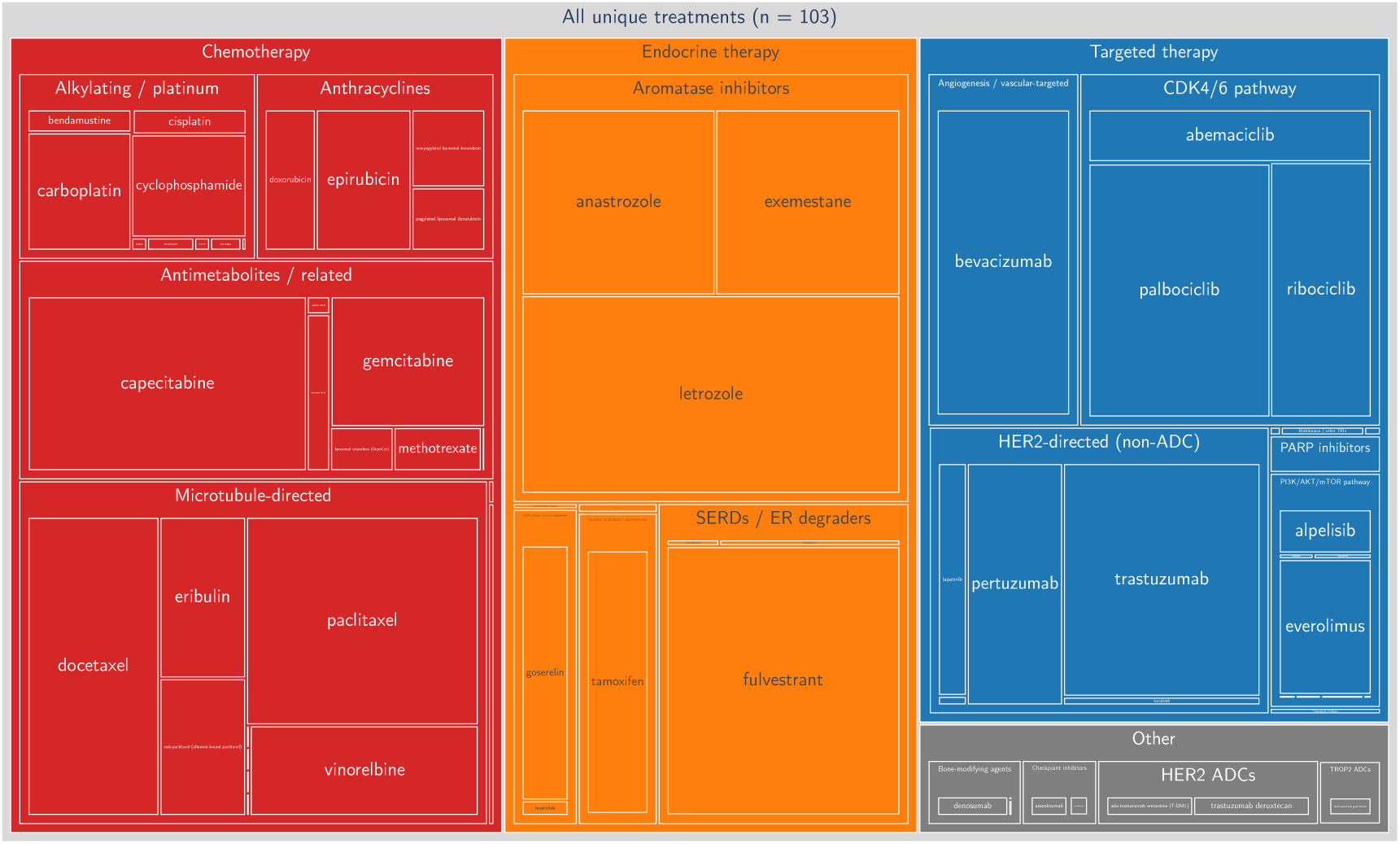
Treemap visualisation of therapies recorded in the AGMT MBC registry. Nested rectangles indicate a nested grouping of treatments, reflecting a hierarchical categorisation. Rectangle size indicates the number of occurrences in the dataset.

### A.2 Visualisation of patient trajectories

**Figure S2:**
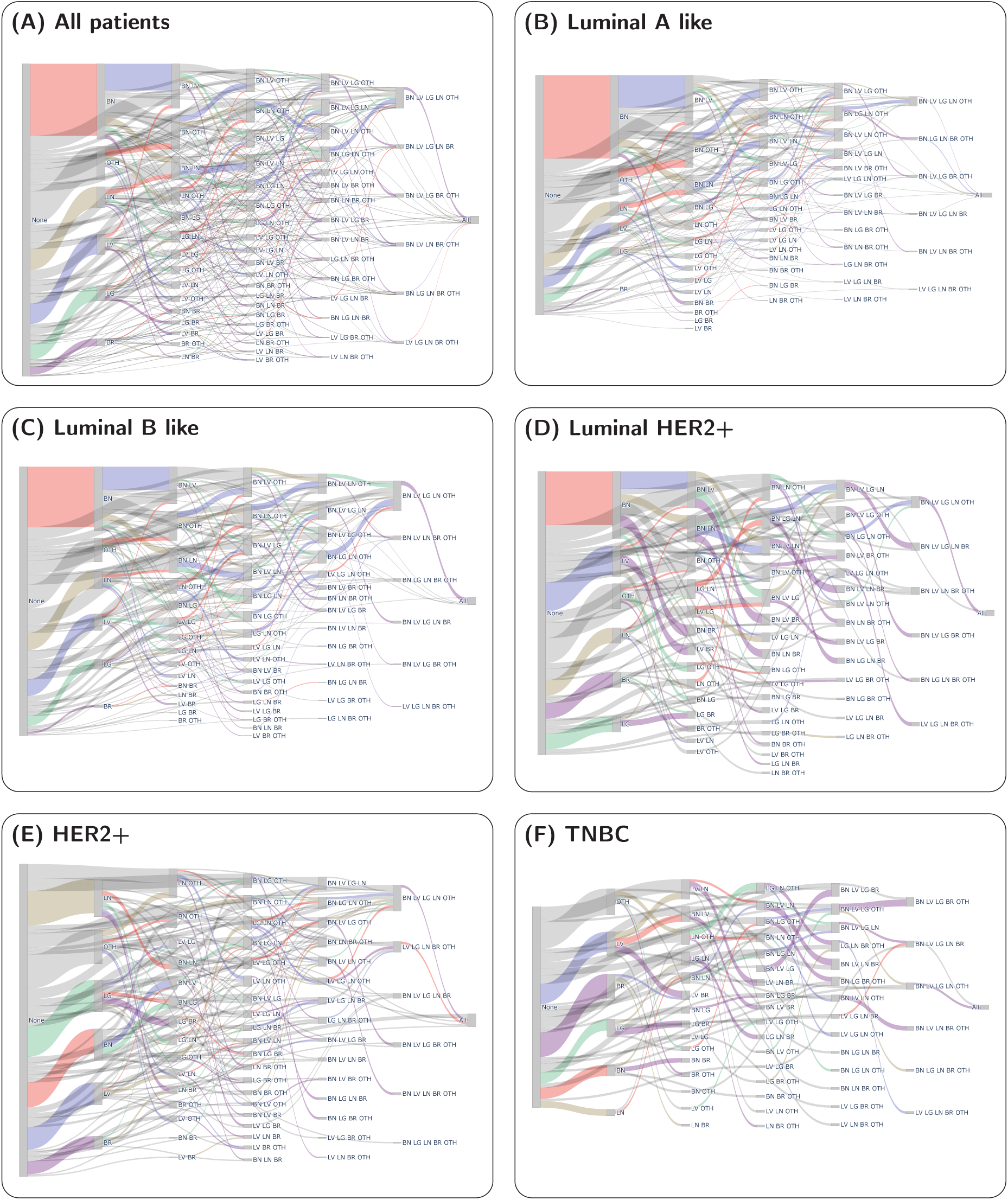
Observed metastasis process for different molecular subtypes. Sankey plots of metastatic progression for different breast cancer subtypes. Each panel displays the flow of metastases between anatomical sites for the indicated subtype. The colour of each direct transition represents the organ added. The abbreviations used represent Bone (BN), Lungs (LG), Liver (LV), Lymph nodes (LN), Brain (BR) and Other (OTH).

## B Mathematical derivation

In this section, we give a more detailed description of the presented framework’s mathematical underpinnings. As outlined above, we consider a set *L* of |*L*| =: *N* organ localisations, and with it a Continuous-Time Markov Chain (CTMC) on the set of all possible configurations {0, 1}*^L^*. We take an enumeration of the resulting 2*^N^* states in topological ordering (i.e. enumerating all *S* ⊆ *L* so that *S_i_* ⊆ *S_j_* =⇒ *i* ≤ *j*) which we identify with (1*, . . .,* 2*^N^*. We now consider non-negative *transition rates* (*q_ij_*)_1≤_*_i,j,_*_≤_*_N_* which may depend on covariate values and model parameters (*x*; *ϑ*), and which we collect into a *generator matrix*

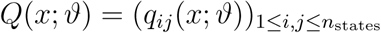

with the convention that the diagonal entries are given as

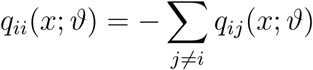

With these assumptions, and given some observation duration *τ*, the CTMC framework then provides a *transition probability matrix P* (*τ, x*; *ϑ*) = (*p_ij_*(*τ, x*; *ϑ*))_1≤_*_i,j_*_≤_*_n_*_states_ where

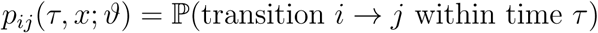

Specifically, they are linked by the matrix exponential relation [Nor98]

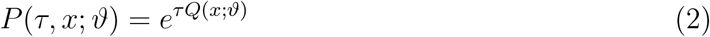

We make a “no self-healing” assumption that metastases do not vanish, so that the generator matrix *Q* becomes upper triangular in our enumeration. For our model, admissible transitions are exactly those which add one organ to an existing metastatic set.

A key property of our Continuous-Time Markov Chain formulation is that equation 2 yields a likelihood function, and thus, in turn – given Bayesian priors on *ϑ* – an un-normalised posterior density. Take a transition *T* = (*i, j, τ, x*) from initial state *i* to final state *j* within time interval *τ*, given the covariate vector *x* updated at the start of the transition interval. Then, we consider the likelihood

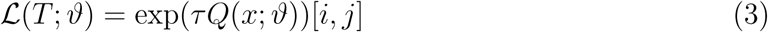

where exp (*τ_k_Q*(*x_k_*; *ϑ*)) refers to the matrix exponential. When considering a collection of transitions, this likelihood is extended by factorisation.

Actually evaluating equation 3 is non-trivial. Applied to a collection of transitions, it necessitates a computation of the matrix exponential – and not just once per objective function evaluation, but once per transition, for potentially several different matrices, and pairwise different values of *τ*. This is computationally prohibitively expensive even for moderate matrix dimensions, and can suffer from numerical issues (see also the seminal *Nineteen dubious ways to compute the exponential of a matrix* by Moler and van Loan [MV78; MV03]).

To make the efficient inference of a model of the size presented here possible, we instead exploit the acyclic topology induced by our choice of state space. As an immediate consequence, it ensures that *Q* is upper triangular, allowing us to use a direct divided-difference identity.

**Definition B.1** (Divided differences, cf. [Hig08], ch. B.16). Let *x*_0_*, x*_1_*, . . ., x_n_* ∈ ℂ be ordered so that equal points are contiguous, that is,

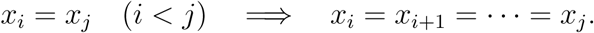

*Divided differences* of a function *f* at the points *x_k_*are defined recursively by

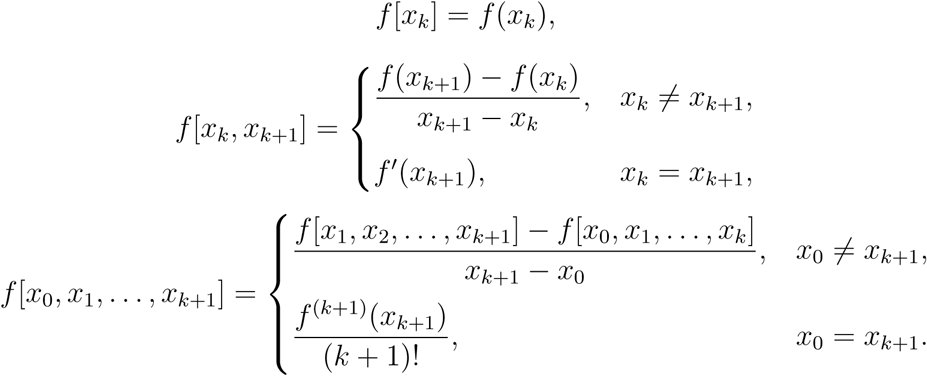

**Theorem B.2** (Function of triangular matrix, cf. [Hig08] Theorem 4.11). Let *Q* ∈ ℂ*^n^*^×^*^n^ be upper triangular and suppose that f is defined on the spectrum of Q. Then F* = *f* (*Q*) *is upper triangular with f_ii_* = *f* (*q_ii_*) *and*

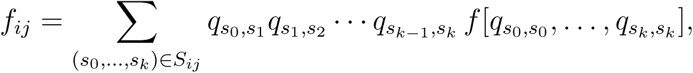

*where λ_i_* = *q_ii_, S_ij_ is the set of all strictly increasing sequences of integers that start at i and end at j*.

Given pairwise different exit rates along paths, this has a closed form (see for example [DD78], ch. 1):

**Lemma B.3.** Let i ≠ *j* =⇒ q_ii_ ≠ *q_jj_. Then,*

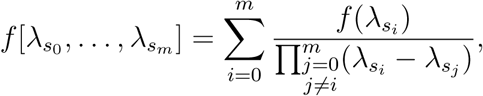

As a consequence, we directly get the formula which we apply numerically, with a pre-computation of the index sequences (*S_ij_*)*_i,j_*:

**Corollary B.4.** Let Q ∈ ℝ*^N×N^* be a rate matrix satisfying the constraint that q_ii_ ≠ q_jj_ whenever i ≠ *j. Then,*

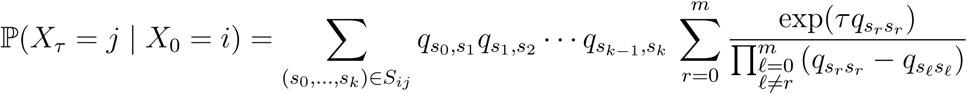

From here, given data consisting of collections of observed transitions, and patients’ covariate values, inference on *ϑ* can then be performed by the maximum a posteriori method for point estimates and Markov Chain Monte Carlo for uncertainty estimation. Moreover, this likelihood formulation is differentiable [NH95], making it possible to efficiently sample from the posterior distribution using Hamiltonian Monte-Carlo [DKP+87].

### B.1 Parametrisation

Let *i* ≡ *S_i_* be an initial state, and consider a transition adding an organ *O*^′^ which results in state *j* ≡ *S_j_* = *S_i_* ∪ {*O*^′^}. Let *x* represent a vector of covariate values. Then, we consider 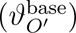*_O′_*_∈_*_L_*, (*β_k_*)*_k_*_=1_*_,…K_* where *K* is determined by the model’s covariate design matrix, and {*γ_O_*_→_*_O′_* | *O* → *O*^′^ is considered}. Together, they make up our model’s actual free parameters *ϑ* = (*ϑ*^base^*, β, γ*), which determine the transition rate matrix *Q*(*x*; *ϑ*) = (*q_ij_*(*x*; *ϑ*))*_i,j_*, where

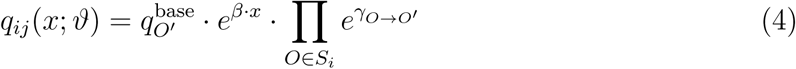

Numerically, we express it in the more stable log-scale

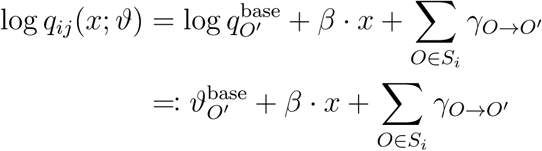

Notably, under *β* ≡ 0*, γ* ≡ 0, the model would reduce to an exponential survival model for time to metastasis detection of the localisations. With non-zero *β*, but *γ* ≡ 0, it can be seen as a coupled collection of exponential baseline accelerated failure time models [Wei92].

This approach of expressing rates of all possible transitions via a small set of biologically meaningful parameters has the advantage that all available data are leveraged, but results can be interpreted very easily at the same time. It stands in contrast to a näıve alternative parametrisation which would be to consider every admissible off-diagonal entry of *Q* a separate parameter, introducing a high number of degrees of freedom with limited identifiability when only few observations of patients in particular states exist.

## C Data processing and modelling details

Trajectory data were processed into transition form. For patients with metachronous metastatic disease, primary breast cancer diagnosis was chosen as the first observation time. For patients who died within the observation period, metastases present at the time of death were included as a final observation. To account for uncertainty in progression order arising from possible delays in documentation, observations were merged when they were strictly less than three months apart, and dated to the later timepoint. As patients underwent evaluation at regular intervals of at most 6 months, we assume that six months before documented metastasis detection, a prior evaluation has taken place. Direct transitions from primary diagnosis to all metastases being present were discarded as non-informative. The resulting transitions, alongside covariate information, were serialised to the inter-operable feather format [Apa16].

For maximum a posteriori estimation, we employed a multistart variant of the `BlackBoxOptim.jl` [FS18] `adaptive_de_rand_1_bin` algorithm implementing differential evolution optimisation [SP97; WLH+14]; with a particle population of 200, and a wall time limit of 12 hours. For each optimisation, we ran 48 starts in parallel.

For HMC sampling, carried out using the AdvancedHMC.jl implementation [XGT+20], we opted for a leapfrog integrator, an adaptive step size, and a target acceptance rate of *δ* = 0.8. We sampled 4 chains in parallel, each consisting of 2000 samples after a burn-in interval of 500.

### C.1 Tabular estimation result details

For each parameter, the tables report posterior summary statistics together with standard Markov chain Monte Carlo convergence diagnostics. The reported *R^* is the modern rank-normalized split-*R*^^^, which compares within-chain and between-chain variation after splitting each chain into halves; values close to 1 indicate good mixing across chains [VGS+21]. We additionally report the classical Gelman–Rubin potential scale reduction factor (PSRF), which quantifies how much the scale of the pooled draws could still decrease if sampling were continued; again, values close to 1 are desirable [GR92; BG98]. The column PSRF_97.5%_ gives the upper 97.5% confidence limit of the PSRF and is therefore a more conservative version of the same diagnostic [BG98]. Finally, ESS_bulk_ and ESS_tail_ denote the effective sample size in the bulk and tails of the posterior distribution, respectively, after accounting for autocorrelation; larger values indicate more efficient sampling, with ESS_bulk_ targeting central posterior behavior and ESS_tail_ targeting estimation of tail quantiles [VGS+21].

**Table S18:** Markov Chain Monte Carlo diagnostics for shared parameters. Detailed estimation results for covariate effects shared across subtypes.

| Parameter | $\hat{R}$ | PSRF | PSRF<sub>97.5%</sub> | ESSb | ESSt | Mean | SD | q2.5 | q25 | q50 | q75 | q97.5 |
| --- | --- | --- | --- | --- | --- | --- | --- | --- | --- | --- | --- | --- |
| Fixed covariate effects |  |  |  |  |  |  |  |  |  |  |  |  |
| Long DFS | 1.003 | 1.002 | 1.006 | 644.978 | 1400.218 | -0.229 | 0.039 | -0.306 | -0.255 | -0.229 | -0.202 | -0.153 |
| Ki67 > 20% | 1.003 | 1.003 | 1.008 | 734.819 | 1189.186 | 0.040 | 0.048 | -0.059 | 0.008 | 0.039 | 0.072 | 0.136 |
| Lobular spectrum | 1.004 | 1.002 | 1.004 | 1133.563 | 1719.529 | -0.153 | 0.058 | -0.266 | -0.193 | -0.153 | -0.115 | -0.043 |
| Unknown histology | 1.002 | 1.003 | 1.008 | 1108.807 | 2017.236 | 0.115 | 0.081 | -0.044 | 0.062 | 0.115 | 0.169 | 0.275 |
| Line of therapy | 1.003 | 1.003 | 1.009 | 1077.737 | 1558.410 | 0.250 | 0.014 | 0.222 | 0.241 | 0.250 | 0.259 | 0.276 |
| Grade 2 | 1.057 | 1.087 | 1.244 | 95.408 | 228.140 | 0.175 | 0.115 | -0.044 | 0.094 | 0.172 | 0.251 | 0.409 |
| Grade 3 | 1.055 | 1.083 | 1.235 | 95.073 | 276.296 | 0.298 | 0.124 | 0.066 | 0.212 | 0.294 | 0.381 | 0.548 |
| Unknown grade | 1.052 | 1.078 | 1.221 | 94.974 | 253.562 | -0.026 | 0.122 | -0.249 | -0.113 | -0.029 | 0.056 | 0.228 |

A visualisation of the estimated posterior distributions on the treatment PC effects can also be found in figure S3.

**Figure S3:**
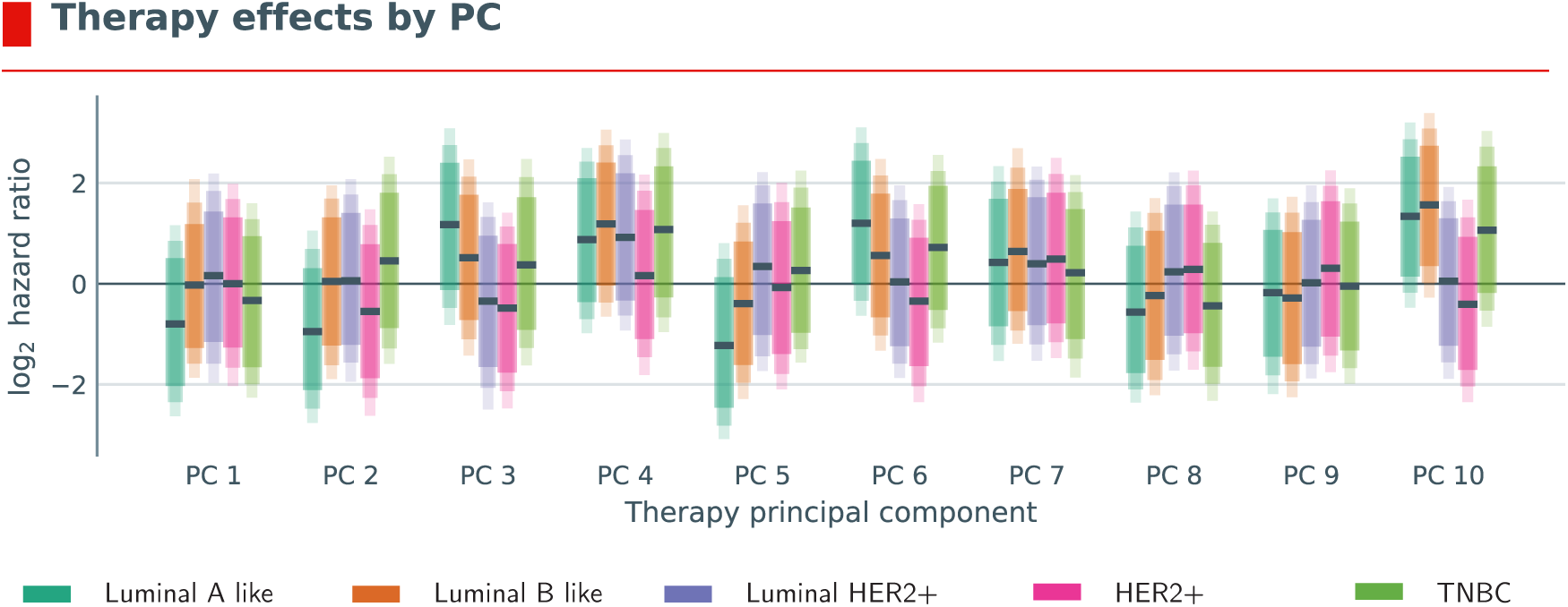
Visualisation of the estimated coefficients (shown with 80%, 90% and 95% credible intervals indicated by shaded areas) for the ten PCA components of the SapBERT embedding of patient therapies. The PCA components themselves are not directly interpretable, but the estimation results suggest that the model has indeed learned to leverage this information, as the coefficients are non-zero and show variation across subtypes.

**Table S19:** Markov Chain Monte Carlo diagnostics for luminal A like disease. Detailed estimation results for parameters relating to the luminal A like group

| Parameter | $\hat{R}$ | PSRF | PSRF<sub>97.5%</sub> | ESSb | ESS<sub>t</sub> | Mean | SD | q2.5 | q25 | q50 | q75 | q97.5 |
| --- | --- | --- | --- | --- | --- | --- | --- | --- | --- | --- | --- | --- |
| Baseline rates |  |  |  |  |  |  |  |  |  |  |  |  |
| log<sub>2</sub> baseline rate Bone | 1.006 | 1.002 | 1.005 | 225.743 | 640.230 | -3.319 | 0.265 | -3.859 | -3.495 | -3.312 | -3.128 | -2.842 |
| log<sub>2</sub> baseline rate Lungs | 1.030 | 1.034 | 1.100 | 186.528 | 433.218 | -4.392 | 0.258 | -4.928 | -4.568 | -4.377 | -4.206 | -3.923 |
| log<sub>2</sub> baseline rate Liver | 1.013 | 1.017 | 1.050 | 192.017 | 417.995 | -4.067 | 0.231 | -4.544 | -4.214 | -4.060 | -3.912 | -3.636 |
| log<sub>2</sub> baseline rate Lymph nodes | 1.029 | 1.019 | 1.058 | 175.644 | 475.173 | -4.027 | 0.266 | -4.573 | -4.199 | -4.015 | -3.845 | -3.536 |
| log<sub>2</sub> baseline rate Brain | 1.026 | 1.036 | 1.106 | 197.058 | 538.973 | -6.091 | 0.332 | -6.792 | -6.295 | -6.079 | -5.872 | -5.464 |
| log<sub>2</sub> baseline rate Other | 1.023 | 1.028 | 1.083 | 235.767 | 786.511 | -4.126 | 0.246 | -4.638 | -4.286 | -4.119 | -3.959 | -3.668 |
| log<sub>2</sub> first metastasis rate Bone | 1.799 | 1.992 | 4.327 | 19.158 | 31.990 | 4.446 | 2.330 | 1.545 | 2.810 | 3.829 | 5.168 | 10.208 |
| log<sub>2</sub> first metastasis rate Lungs | 1.808 | 1.997 | 4.260 | 19.303 | 32.664 | 2.489 | 2.343 | -0.404 | 0.808 | 1.901 | 3.232 | 8.331 |
| log<sub>2</sub> first metastasis rate Liver | 1.772 | 1.973 | 4.219 | 19.287 | 31.603 | 2.472 | 2.333 | -0.461 | 0.844 | 1.851 | 3.198 | 8.244 |
| log<sub>2</sub> first metastasis rate Lymph nodes | 1.776 | 1.979 | 4.279 | 19.317 | 31.791 | 2.829 | 2.333 | -0.097 | 1.203 | 2.211 | 3.519 | 8.572 |
| log<sub>2</sub> first metastasis rate Brain | 1.651 | 1.898 | 3.745 | 21.342 | 34.510 | -0.169 | 2.427 | -3.495 | -1.792 | -0.748 | 0.723 | 5.952 |
| log<sub>2</sub> first metastasis rate Other | 1.791 | 1.987 | 4.301 | 19.202 | 31.547 | 3.384 | 2.332 | 0.512 | 1.743 | 2.760 | 4.089 | 9.136 |
| Fixed covariate effects |  |  |  |  |  |  |  |  |  |  |  |  |
| Pre-menopausal | 1.010 | 1.009 | 1.022 | 317.130 | 1012.175 | -0.081 | 0.136 | -0.351 | -0.171 | -0.079 | 0.014 | 0.175 |
| Unknown menopausal status | 1.009 | 1.003 | 1.007 | 816.761 | 1397.545 | 0.120 | 0.090 | -0.055 | 0.057 | 0.120 | 0.181 | 0.298 |
| Age (pre-menopausal) | 1.012 | 1.009 | 1.026 | 350.448 | 878.843 | -0.753 | 0.634 | -1.976 | -1.176 | -0.741 | -0.322 | 0.466 |
| Age (unknown menopausal status) | 1.008 | 1.002 | 1.007 | 663.092 | 972.712 | 0.318 | 0.597 | -0.874 | -0.088 | 0.322 | 0.732 | 1.458 |
| Age (post-menopausal) | 1.022 | 1.009 | 1.027 | 455.199 | 660.797 | -1.078 | 0.372 | -1.821 | -1.322 | -1.075 | -0.829 | -0.357 |
| Therapy embedding effects |  |  |  |  |  |  |  |  |  |  |  |  |
| Therapy PC 1 | 1.001 | 1.001 | 1.003 | 1026.986 | 1586.987 | -0.543 | 0.682 | -1.822 | -0.996 | -0.554 | -0.094 | 0.806 |
| Therapy PC 2 | 1.003 | 1.002 | 1.006 | 545.279 | 1006.347 | -0.634 | 0.656 | -1.915 | -1.070 | -0.657 | -0.199 | 0.732 |
| Therapy PC 3 | 1.002 | 1.003 | 1.007 | 1239.900 | 1680.874 | 0.800 | 0.686 | -0.565 | 0.355 | 0.814 | 1.252 | 2.140 |
| Therapy PC 4 | 1.003 | 1.002 | 1.006 | 1113.786 | 1515.237 | 0.604 | 0.657 | -0.681 | 0.163 | 0.609 | 1.045 | 1.867 |
| Therapy PC 5 | 1.004 | 1.004 | 1.010 | 945.112 | 1602.559 | -0.827 | 0.698 | -2.136 | -1.304 | -0.850 | -0.353 | 0.558 |
| Therapy PC 6 | 1.006 | 1.005 | 1.008 | 632.670 | 1200.955 | 0.837 | 0.661 | -0.439 | 0.394 | 0.833 | 1.288 | 2.152 |
| Therapy PC 7 | 1.016 | 1.015 | 1.042 | 426.098 | 1017.940 | 0.289 | 0.686 | -1.060 | -0.177 | 0.293 | 0.758 | 1.619 |
| Therapy PC 8 | 1.008 | 1.006 | 1.013 | 649.560 | 1140.791 | -0.369 | 0.684 | -1.634 | -0.855 | -0.390 | 0.103 | 0.994 |
| Therapy PC 9 | 1.002 | 1.002 | 1.005 | 985.128 | 1508.516 | -0.134 | 0.686 | -1.516 | -0.608 | -0.122 | 0.339 | 1.174 |
| Therapy PC 10 | 1.006 | 1.002 | 1.007 | 1295.016 | 2142.708 | 0.924 | 0.648 | -0.328 | 0.482 | 0.929 | 1.352 | 2.220 |
| Interaction effects |  |  |  |  |  |  |  |  |  |  |  |  |
| Lungs → Bone interaction | 1.003 | 1.001 | 1.004 | 895.163 | 1895.336 | 0.034 | 0.220 | -0.398 | -0.116 | 0.038 | 0.186 | 0.455 |
| Liver → Bone interaction | 1.004 | 1.004 | 1.010 | 929.385 | 2083.286 | 0.252 | 0.239 | -0.221 | 0.091 | 0.250 | 0.413 | 0.716 |
| Lymph nodes → Bone interaction | 1.008 | 1.009 | 1.029 | 522.618 | 815.858 | 0.096 | 0.212 | -0.308 | -0.048 | 0.096 | 0.236 | 0.526 |
| Other → Bone interaction | 1.007 | 1.010 | 1.028 | 439.041 | 778.153 | 0.157 | 0.229 | -0.263 | 0.001 | 0.146 | 0.309 | 0.636 |
| Bone → Lungs interaction | 1.007 | 1.005 | 1.013 | 431.047 | 610.821 | 0.176 | 0.216 | -0.223 | 0.028 | 0.169 | 0.314 | 0.623 |
| Liver → Lungs interaction | 1.013 | 1.008 | 1.024 | 396.382 | 741.158 | 0.722 | 0.212 | 0.301 | 0.581 | 0.723 | 0.866 | 1.135 |
| Lymph nodes → Lungs interaction | 1.010 | 1.012 | 1.036 | 482.587 | 1074.979 | 0.566 | 0.209 | 0.136 | 0.430 | 0.572 | 0.708 | 0.972 |
| Other → Lungs interaction | 1.006 | 1.004 | 1.011 | 631.918 | 1065.110 | 0.130 | 0.219 | -0.295 | -0.015 | 0.126 | 0.276 | 0.563 |
| Bone → Liver interaction | 1.002 | 1.001 | 1.003 | 334.973 | 465.368 | 1.166 | 0.193 | 0.811 | 1.028 | 1.161 | 1.289 | 1.581 |
| Lungs → Liver interaction | 1.008 | 1.009 | 1.026 | 394.732 | 940.154 | 0.155 | 0.179 | -0.200 | 0.034 | 0.160 | 0.280 | 0.502 |
| Lymph nodes → Liver interaction | 1.005 | 1.006 | 1.016 | 599.600 | 920.352 | 0.078 | 0.159 | -0.244 | -0.026 | 0.081 | 0.190 | 0.369 |
| Other → Liver interaction | 1.007 | 1.009 | 1.026 | 447.381 | 911.081 | -0.089 | 0.156 | -0.393 | -0.195 | -0.088 | 0.015 | 0.215 |
| Bone → Lymph nodes interaction | 1.023 | 1.015 | 1.046 | 457.266 | 927.014 | 0.087 | 0.217 | -0.326 | -0.059 | 0.085 | 0.228 | 0.516 |
| Lungs → Lymph nodes interaction | 1.009 | 1.014 | 1.039 | 340.857 | 686.408 | 0.277 | 0.254 | -0.233 | 0.111 | 0.285 | 0.447 | 0.759 |
| Liver → Lymph nodes interaction | 1.006 | 1.008 | 1.024 | 818.164 | 1691.511 | 0.251 | 0.216 | -0.162 | 0.104 | 0.247 | 0.400 | 0.667 |
| Other → Lymph nodes interaction | 1.016 | 1.007 | 1.021 | 476.606 | 874.625 | 0.189 | 0.223 | -0.265 | 0.045 | 0.194 | 0.338 | 0.610 |
| Bone → Brain interaction | 1.011 | 1.020 | 1.055 | 424.570 | 691.409 | 0.702 | 0.294 | 0.144 | 0.503 | 0.693 | 0.888 | 1.315 |
| Lungs → Brain interaction | 1.003 | 1.002 | 1.006 | 831.510 | 1385.959 | 0.369 | 0.247 | -0.119 | 0.202 | 0.378 | 0.538 | 0.831 |
| Liver → Brain interaction | 1.003 | 1.003 | 1.010 | 1227.839 | 1938.325 | 0.810 | 0.244 | 0.333 | 0.645 | 0.807 | 0.974 | 1.294 |
| Lymph nodes → Brain interaction | 1.020 | 1.007 | 1.020 | 680.314 | 994.248 | 0.262 | 0.243 | -0.222 | 0.098 | 0.264 | 0.428 | 0.722 |
| Other → Brain interaction | 1.005 | 1.002 | 1.004 | 509.505 | 1074.566 | 0.498 | 0.261 | -0.010 | 0.320 | 0.493 | 0.677 | 0.997 |
| Bone → Other interaction | 1.003 | 1.002 | 1.008 | 737.304 | 1306.409 | 0.634 | 0.204 | 0.251 | 0.494 | 0.627 | 0.766 | 1.057 |
| Lungs → Other interaction | 1.009 | 1.004 | 1.010 | 409.064 | 1152.468 | 0.518 | 0.180 | 0.159 | 0.395 | 0.520 | 0.643 | 0.862 |
| Liver → Other interaction | 1.006 | 1.001 | 1.004 | 655.564 | 1636.056 | 0.414 | 0.158 | 0.103 | 0.309 | 0.411 | 0.520 | 0.726 |
| Lymph nodes → Other interaction | 1.001 | 1.001 | 1.004 | 1012.646 | 1613.091 | 0.471 | 0.167 | 0.137 | 0.360 | 0.472 | 0.583 | 0.799 |

**Table S20:** Markov Chain Monte Carlo diagnostics for luminal B like disease. Detailed estimation results for parameters relating to the luminal B like group

| Parameter | $\hat{R}$ | PSRF | PSRF<sub>97.5%</sub> | ESSb | ESS<sub>t</sub> | Mean | SD | q2.5 | q25 | q50 | q75 | q97.5 |
| --- | --- | --- | --- | --- | --- | --- | --- | --- | --- | --- | --- | --- |
| Baseline rates |  |  |  |  |  |  |  |  |  |  |  |  |
| log<sub>2</sub> baseline rate Bone | 1.040 | 1.051 | 1.147 | 157.241 | 458.564 | -3.435 | 0.282 | -3.995 | -3.622 | -3.437 | -3.240 | -2.887 |
| log<sub>2</sub> baseline rate Lungs | 1.017 | 1.023 | 1.059 | 246.043 | 638.055 | -3.936 | 0.249 | -4.431 | -4.105 | -3.929 | -3.759 | -3.478 |
| log<sub>2</sub> baseline rate Liver | 1.021 | 1.031 | 1.093 | 154.526 | 421.266 | -3.974 | 0.233 | -4.446 | -4.132 | -3.972 | -3.814 | -3.531 |
| log<sub>2</sub> baseline rate Lymph nodes | 1.014 | 1.024 | 1.063 | 215.588 | 476.683 | -3.543 | 0.268 | -4.056 | -3.724 | -3.543 | -3.367 | -3.011 |
| log<sub>2</sub> baseline rate Brain | 1.011 | 1.012 | 1.035 | 281.940 | 936.386 | -5.652 | 0.314 | -6.298 | -5.858 | -5.645 | -5.436 | -5.051 |
| log<sub>2</sub> baseline rate Other | 1.028 | 1.044 | 1.125 | 226.763 | 757.068 | -3.928 | 0.263 | -4.482 | -4.100 | -3.912 | -3.743 | -3.441 |
| log<sub>2</sub> first metastasis rate Bone | 1.807 | 1.995 | 3.382 | 19.508 | 35.390 | 4.608 | 2.105 | 1.478 | 3.053 | 4.308 | 5.530 | 9.618 |
| log<sub>2</sub> first metastasis rate Lungs | 1.799 | 1.996 | 3.378 | 19.638 | 34.929 | 3.162 | 2.108 | -0.004 | 1.641 | 2.897 | 4.096 | 8.156 |
| log<sub>2</sub> first metastasis rate Liver | 1.789 | 1.992 | 3.368 | 19.626 | 34.927 | 3.264 | 2.115 | 0.127 | 1.708 | 3.001 | 4.203 | 8.255 |
| log<sub>2</sub> first metastasis rate Lymph nodes | 1.797 | 1.987 | 3.358 | 19.565 | 35.493 | 3.606 | 2.116 | 0.419 | 2.025 | 3.352 | 4.550 | 8.664 |
| log<sub>2</sub> first metastasis rate Brain | 1.699 | 1.843 | 2.952 | 20.828 | 37.450 | 0.539 | 2.161 | -2.897 | -1.040 | 0.256 | 1.681 | 5.640 |
| log<sub>2</sub> first metastasis rate Other | 1.805 | 2.002 | 3.397 | 19.539 | 34.407 | 3.689 | 2.107 | 0.528 | 2.139 | 3.423 | 4.614 | 8.703 |
| Fixed covariate effects |  |  |  |  |  |  |  |  |  |  |  |  |
| Pre-menopausal | 1.004 | 1.001 | 1.002 | 583.478 | 1181.897 | 0.439 | 0.146 | 0.146 | 0.342 | 0.442 | 0.538 | 0.721 |
| Unknown menopausal status | 1.006 | 1.009 | 1.026 | 614.751 | 1365.456 | 0.189 | 0.099 | -0.009 | 0.123 | 0.190 | 0.256 | 0.377 |
| Age (pre-menopausal) | 1.005 | 1.003 | 1.008 | 587.365 | 1196.461 | -0.317 | 0.600 | -1.490 | -0.709 | -0.314 | 0.081 | 0.838 |
| Age (unknown menopausal status) | 1.014 | 1.022 | 1.065 | 452.075 | 1092.752 | -1.037 | 0.550 | -2.069 | -1.421 | -1.047 | -0.659 | 0.065 |
| Age (post-menopausal) | 1.005 | 1.004 | 1.010 | 483.078 | 758.194 | -0.888 | 0.379 | -1.624 | -1.137 | -0.890 | -0.638 | -0.128 |
| Therapy embedding effects |  |  |  |  |  |  |  |  |  |  |  |  |
| Therapy PC 1 | 1.013 | 1.020 | 1.061 | 266.792 | 625.691 | -0.014 | 0.675 | -1.293 | -0.463 | -0.017 | 0.423 | 1.441 |
| Therapy PC 2 | 1.001 | 1.001 | 1.004 | 2490.313 | 2901.816 | 0.029 | 0.680 | -1.310 | -0.418 | 0.033 | 0.480 | 1.354 |
| Therapy PC 3 | 1.003 | 1.004 | 1.011 | 884.494 | 1661.230 | 0.361 | 0.684 | -0.988 | -0.096 | 0.358 | 0.820 | 1.710 |
| Therapy PC 4 | 1.003 | 1.001 | 1.002 | 590.509 | 1401.609 | 0.824 | 0.656 | -0.453 | 0.381 | 0.822 | 1.267 | 2.121 |
| Therapy PC 5 | 1.007 | 1.006 | 1.014 | 836.160 | 1033.417 | -0.263 | 0.662 | -1.585 | -0.688 | -0.272 | 0.167 | 1.079 |
| Therapy PC 6 | 1.004 | 1.001 | 1.002 | 1621.773 | 2180.519 | 0.391 | 0.667 | -0.921 | -0.036 | 0.389 | 0.833 | 1.718 |
| Therapy PC 7 | 1.012 | 1.012 | 1.029 | 551.153 | 533.843 | 0.462 | 0.668 | -0.826 | 0.032 | 0.444 | 0.882 | 1.866 |
| Therapy PC 8 | 1.002 | 1.003 | 1.008 | 1190.180 | 2239.634 | -0.164 | 0.694 | -1.532 | -0.624 | -0.161 | 0.307 | 1.177 |
| Therapy PC 9 | 1.006 | 1.001 | 1.001 | 724.546 | 1067.110 | -0.192 | 0.706 | -1.559 | -0.667 | -0.197 | 0.284 | 1.195 |
| Therapy PC 10 | 1.002 | 1.001 | 1.003 | 1345.561 | 2201.819 | 1.082 | 0.640 | -0.191 | 0.659 | 1.085 | 1.511 | 2.348 |
| Interaction effects |  |  |  |  |  |  |  |  |  |  |  |  |
| Lungs → Bone interaction | 1.003 | 1.002 | 1.005 | 690.528 | 1256.053 | 0.289 | 0.227 | -0.164 | 0.140 | 0.293 | 0.444 | 0.723 |
| Liver → Bone interaction | 1.011 | 1.010 | 1.031 | 588.249 | 1692.005 | 0.632 | 0.244 | 0.160 | 0.469 | 0.628 | 0.799 | 1.107 |
| Lymph nodes → Bone interaction | 1.025 | 1.030 | 1.084 | 362.767 | 856.825 | 0.589 | 0.218 | 0.172 | 0.439 | 0.582 | 0.729 | 1.037 |
| Other → Bone interaction | 1.004 | 1.001 | 1.003 | 836.852 | 1215.626 | -0.152 | 0.228 | -0.582 | -0.310 | -0.157 | 0.005 | 0.294 |
| Bone → Lungs interaction | 1.023 | 1.026 | 1.076 | 360.766 | 606.011 | -0.168 | 0.212 | -0.572 | -0.310 | -0.174 | -0.023 | 0.261 |
| Liver → Lungs interaction | 1.004 | 1.005 | 1.014 | 793.725 | 1421.589 | 0.433 | 0.230 | -0.035 | 0.279 | 0.438 | 0.593 | 0.867 |
| Lymph nodes → Lungs interaction | 1.003 | 1.001 | 1.003 | 785.543 | 1526.238 | 0.551 | 0.180 | 0.199 | 0.429 | 0.552 | 0.673 | 0.907 |
| Other → Lungs interaction | 1.003 | 1.004 | 1.012 | 688.964 | 1092.599 | 0.268 | 0.190 | -0.102 | 0.141 | 0.265 | 0.395 | 0.652 |
| Bone → Liver interaction | 1.008 | 1.012 | 1.036 | 324.700 | 749.132 | 0.775 | 0.180 | 0.426 | 0.655 | 0.773 | 0.890 | 1.129 |
| Lungs → Liver interaction | 1.009 | 1.003 | 1.007 | 668.859 | 1472.552 | 0.566 | 0.179 | 0.206 | 0.448 | 0.572 | 0.686 | 0.912 |
| Lymph nodes → Liver interaction | 1.003 | 1.002 | 1.005 | 827.068 | 1304.428 | 0.161 | 0.146 | -0.124 | 0.058 | 0.160 | 0.259 | 0.455 |
| Other → Liver interaction | 1.003 | 1.003 | 1.006 | 874.251 | 1391.086 | 0.308 | 0.151 | 0.007 | 0.207 | 0.303 | 0.413 | 0.598 |
| Bone → Lymph nodes interaction | 1.002 | 1.005 | 1.011 | 603.678 | 1039.351 | -0.254 | 0.219 | -0.670 | -0.403 | -0.258 | -0.111 | 0.193 |
| Lungs → Lymph nodes interaction | 1.003 | 1.001 | 1.004 | 931.185 | 1974.861 | 0.449 | 0.220 | 0.022 | 0.300 | 0.451 | 0.598 | 0.876 |
| Liver → Lymph nodes interaction | 1.003 | 1.005 | 1.014 | 837.149 | 1473.309 | -0.189 | 0.220 | -0.627 | -0.333 | -0.189 | -0.042 | 0.233 |
| Other → Lymph nodes interaction | 1.003 | 1.004 | 1.009 | 835.000 | 1596.940 | 0.164 | 0.208 | -0.264 | 0.027 | 0.171 | 0.306 | 0.559 |
| Bone → Brain interaction | 1.004 | 1.003 | 1.007 | 632.529 | 965.791 | 0.065 | 0.277 | -0.454 | -0.127 | 0.061 | 0.247 | 0.595 |
| Lungs → Brain interaction | 1.001 | 1.000 | 1.001 | 778.312 | 1643.528 | 0.614 | 0.240 | 0.162 | 0.445 | 0.610 | 0.785 | 1.072 |
| Liver → Brain interaction | 1.002 | 1.004 | 1.009 | 914.080 | 1443.408 | 0.823 | 0.248 | 0.333 | 0.656 | 0.819 | 0.990 | 1.311 |
| Lymph nodes → Brain interaction | 1.002 | 1.001 | 1.002 | 775.306 | 1230.035 | 0.223 | 0.254 | -0.286 | 0.055 | 0.223 | 0.399 | 0.702 |
| Other → Brain interaction | 1.005 | 1.004 | 1.013 | 479.648 | 1005.670 | 0.600 | 0.247 | 0.110 | 0.432 | 0.598 | 0.777 | 1.069 |
| Bone → Other interaction | 1.010 | 1.009 | 1.024 | 567.616 | 1065.695 | 0.544 | 0.227 | 0.144 | 0.381 | 0.530 | 0.691 | 1.021 |
| Lungs → Other interaction | 1.006 | 1.003 | 1.007 | 765.304 | 1120.948 | 0.159 | 0.190 | -0.224 | 0.034 | 0.159 | 0.285 | 0.529 |
| Liver → Other interaction | 1.005 | 1.003 | 1.008 | 663.211 | 1128.566 | 0.222 | 0.174 | -0.112 | 0.106 | 0.221 | 0.336 | 0.561 |
| Lymph nodes → Other interaction | 1.005 | 1.004 | 1.013 | 1070.977 | 1955.977 | 0.401 | 0.163 | 0.067 | 0.294 | 0.404 | 0.510 | 0.712 |

**Table S21:** Markov Chain Monte Carlo diagnostics for luminal HER2+ disease. Detailed estimation results for parameters relating to the luminal HER2+ group

| Parameter | $\hat{R}$ | PSRF | PSRF<sub>97.5%</sub> | ESSb | ESS<sub>t</sub> | Mean | SD | q2.5 | q25 | q50 | q75 | q97.5 |
| --- | --- | --- | --- | --- | --- | --- | --- | --- | --- | --- | --- | --- |
| Baseline rates |  |  |  |  |  |  |  |  |  |  |  |  |
| log<sub>2</sub> baseline rate Bone | 1.004 | 1.007 | 1.019 | 501.412 | 1197.614 | -3.447 | 0.355 | -4.162 | -3.692 | -3.433 | -3.204 | -2.782 |
| log<sub>2</sub> baseline rate Lungs | 1.040 | 1.041 | 1.119 | 180.757 | 351.506 | -3.238 | 0.333 | -3.901 | -3.461 | -3.238 | -3.021 | -2.581 |
| log<sub>2</sub> baseline rate Liver | 1.012 | 1.010 | 1.028 | 217.183 | 653.311 | -4.344 | 0.387 | -5.129 | -4.601 | -4.336 | -4.085 | -3.601 |
| log<sub>2</sub> baseline rate Lymph nodes | 1.009 | 1.001 | 1.002 | 350.755 | 897.406 | -4.094 | 0.369 | -4.835 | -4.343 | -4.079 | -3.837 | -3.417 |
| log<sub>2</sub> baseline rate Brain | 1.013 | 1.019 | 1.051 | 256.161 | 493.447 | -4.477 | 0.317 | -5.133 | -4.681 | -4.469 | -4.263 | -3.875 |
| log<sub>2</sub> baseline rate Other | 1.021 | 1.011 | 1.032 | 333.890 | 795.658 | -3.790 | 0.318 | -4.413 | -4.003 | -3.783 | -3.570 | -3.198 |
| log<sub>2</sub> first metastasis rate Bone | 2.182 | 2.433 | 4.938 | 18.594 | 28.421 | 6.173 | 3.876 | 0.823 | 3.364 | 4.811 | 8.737 | 14.837 |
| log<sub>2</sub> first metastasis rate Lungs | 2.224 | 2.456 | 4.963 | 18.495 | 28.704 | 5.263 | 3.908 | -0.220 | 2.436 | 3.940 | 7.820 | 14.022 |
| log<sub>2</sub> first metastasis rate Liver | 2.173 | 2.421 | 4.852 | 18.622 | 29.147 | 5.495 | 3.885 | 0.001 | 2.660 | 4.128 | 8.127 | 14.171 |
| log<sub>2</sub> first metastasis rate Lymph nodes | 2.121 | 2.397 | 4.809 | 18.791 | 29.295 | 5.052 | 3.876 | -0.272 | 2.269 | 3.666 | 7.733 | 13.744 |
| log<sub>2</sub> first metastasis rate Brain | 2.152 | 2.425 | 4.886 | 18.732 | 28.197 | 4.369 | 3.901 | -0.950 | 1.534 | 3.015 | 6.955 | 13.008 |
| log<sub>2</sub> first metastasis rate Other | 2.187 | 2.441 | 4.940 | 18.567 | 28.357 | 5.196 | 3.876 | -0.213 | 2.443 | 3.831 | 7.742 | 13.909 |
| Fixed covariate effects |  |  |  |  |  |  |  |  |  |  |  |  |
| Pre-menopausal | 1.004 | 1.001 | 1.002 | 847.380 | 1395.506 | -0.114 | 0.204 | -0.510 | -0.249 | -0.118 | 0.019 | 0.295 |
| Unknown menopausal status | 1.017 | 1.009 | 1.026 | 480.496 | 877.367 | 0.149 | 0.158 | -0.166 | 0.042 | 0.150 | 0.255 | 0.455 |
| Age (pre-menopausal) | 1.002 | 1.001 | 1.002 | 1162.617 | 2456.142 | -0.558 | 0.639 | -1.810 | -0.987 | -0.551 | -0.135 | 0.679 |
| Age (unknown menopausal status) | 1.003 | 1.001 | 1.002 | 470.081 | 929.240 | 0.248 | 0.649 | -1.017 | -0.178 | 0.245 | 0.694 | 1.490 |
| Age (post-menopausal) | 1.009 | 1.002 | 1.006 | 673.639 | 1619.080 | -0.100 | 0.542 | -1.188 | -0.462 | -0.097 | 0.268 | 0.953 |
| Therapy embedding effects |  |  |  |  |  |  |  |  |  |  |  |  |
| Therapy PC 1 | 1.003 | 1.002 | 1.004 | 510.244 | 1053.363 | 0.103 | 0.710 | -1.364 | -0.347 | 0.111 | 0.571 | 1.517 |
| Therapy PC 2 | 1.006 | 1.003 | 1.006 | 782.427 | 1351.880 | 0.048 | 0.711 | -1.350 | -0.440 | 0.040 | 0.522 | 1.439 |
| Therapy PC 3 | 1.006 | 1.007 | 1.016 | 454.697 | 771.998 | -0.241 | 0.713 | -1.732 | -0.695 | -0.238 | 0.241 | 1.118 |
| Therapy PC 4 | 1.004 | 1.002 | 1.004 | 576.880 | 1029.600 | 0.642 | 0.681 | -0.644 | 0.166 | 0.637 | 1.099 | 1.985 |
| Therapy PC 5 | 1.022 | 1.002 | 1.004 | 255.654 | 696.950 | 0.219 | 0.700 | -1.201 | -0.233 | 0.238 | 0.704 | 1.534 |
| Therapy PC 6 | 1.003 | 1.004 | 1.012 | 481.154 | 838.934 | 0.019 | 0.686 | -1.296 | -0.445 | 0.025 | 0.471 | 1.353 |
| Therapy PC 7 | 1.007 | 1.002 | 1.004 | 875.139 | 1642.016 | 0.284 | 0.686 | -1.059 | -0.186 | 0.277 | 0.734 | 1.613 |
| Therapy PC 8 | 1.003 | 1.000 | 1.001 | 880.512 | 1454.311 | 0.179 | 0.700 | -1.193 | -0.295 | 0.165 | 0.644 | 1.531 |
| Therapy PC 9 | 1.018 | 1.022 | 1.066 | 318.919 | 849.524 | 0.009 | 0.672 | -1.302 | -0.443 | 0.013 | 0.444 | 1.352 |
| Therapy PC 10 | 1.011 | 1.005 | 1.016 | 695.035 | 1331.938 | 0.029 | 0.679 | -1.309 | -0.432 | 0.035 | 0.488 | 1.331 |
| Interaction effects |  |  |  |  |  |  |  |  |  |  |  |  |
| Lungs → Bone interaction | 1.003 | 1.004 | 1.012 | 869.817 | 1708.176 | 0.032 | 0.367 | -0.702 | -0.213 | 0.050 | 0.282 | 0.716 |
| Liver → Bone interaction | 1.002 | 1.003 | 1.008 | 606.237 | 1217.663 | -0.081 | 0.365 | -0.795 | -0.324 | -0.076 | 0.162 | 0.625 |
| Lymph nodes → Bone interaction | 1.010 | 1.015 | 1.046 | 332.240 | 604.470 | -0.163 | 0.400 | -0.966 | -0.430 | -0.163 | 0.116 | 0.601 |
| Other → Bone interaction | 1.012 | 1.007 | 1.020 | 630.945 | 1099.083 | -0.059 | 0.383 | -0.845 | -0.316 | -0.057 | 0.204 | 0.655 |
| Bone → Lungs interaction | 1.016 | 1.019 | 1.051 | 391.386 | 1054.545 | -0.613 | 0.297 | -1.194 | -0.812 | -0.614 | -0.417 | -0.026 |
| Liver → Lungs interaction | 1.020 | 1.026 | 1.076 | 205.284 | 291.953 | -0.121 | 0.318 | -0.736 | -0.330 | -0.117 | 0.088 | 0.497 |
| Lymph nodes → Lungs interaction | 1.004 | 1.003 | 1.008 | 709.089 | 1395.268 | -0.265 | 0.358 | -0.984 | -0.492 | -0.261 | -0.027 | 0.428 |
| Other → Lungs interaction | 1.004 | 1.005 | 1.015 | 705.132 | 1476.702 | 0.099 | 0.323 | -0.529 | -0.116 | 0.101 | 0.314 | 0.733 |
| Bone → Liver interaction | 1.006 | 1.007 | 1.019 | 333.369 | 630.913 | 0.919 | 0.353 | 0.220 | 0.690 | 0.927 | 1.152 | 1.605 |
| Lungs → Liver interaction | 1.002 | 1.004 | 1.011 | 626.263 | 1216.636 | 0.287 | 0.338 | -0.382 | 0.062 | 0.292 | 0.520 | 0.943 |
| Lymph nodes → Liver interaction | 1.009 | 1.004 | 1.012 | 557.699 | 891.312 | -0.148 | 0.325 | -0.776 | -0.367 | -0.147 | 0.073 | 0.496 |
| Other → Liver interaction | 1.007 | 1.004 | 1.012 | 1034.845 | 1765.149 | -0.202 | 0.324 | -0.843 | -0.422 | -0.199 | 0.021 | 0.429 |
| Bone → Lymph nodes interaction | 1.008 | 1.006 | 1.015 | 434.617 | 702.587 | 0.236 | 0.353 | -0.434 | -0.003 | 0.231 | 0.469 | 0.941 |
| Lungs → Lymph nodes interaction | 1.011 | 1.008 | 1.019 | 378.862 | 1042.329 | 0.912 | 0.306 | 0.288 | 0.722 | 0.918 | 1.115 | 1.496 |
| Liver → Lymph nodes interaction | 1.003 | 1.002 | 1.004 | 818.209 | 1584.097 | -0.128 | 0.313 | -0.751 | -0.341 | -0.123 | 0.088 | 0.470 |
| Other → Lymph nodes interaction | 1.002 | 1.001 | 1.003 | 738.219 | 1204.456 | -0.349 | 0.380 | -1.119 | -0.595 | -0.341 | -0.091 | 0.372 |
| Bone → Brain interaction | 1.004 | 1.004 | 1.007 | 778.714 | 1334.667 | 0.505 | 0.253 | 0.017 | 0.334 | 0.497 | 0.667 | 1.020 |
| Lungs → Brain interaction | 1.009 | 1.001 | 1.005 | 570.685 | 1317.018 | 0.208 | 0.260 | -0.313 | 0.039 | 0.208 | 0.384 | 0.709 |
| Liver → Brain interaction | 1.008 | 1.008 | 1.013 | 828.448 | 1902.511 | 0.774 | 0.226 | 0.346 | 0.624 | 0.766 | 0.925 | 1.220 |
| Lymph nodes → Brain interaction | 1.008 | 1.002 | 1.004 | 607.789 | 1148.518 | 0.135 | 0.254 | -0.360 | -0.039 | 0.140 | 0.304 | 0.630 |
| Other → Brain interaction | 1.001 | 1.000 | 1.001 | 1108.651 | 1545.440 | -0.406 | 0.259 | -0.925 | -0.577 | -0.405 | -0.235 | 0.104 |
| Bone → Other interaction | 1.012 | 1.004 | 1.012 | 563.078 | 1366.473 | 0.338 | 0.274 | -0.187 | 0.144 | 0.338 | 0.526 | 0.874 |
| Lungs → Other interaction | 1.003 | 1.003 | 1.006 | 729.239 | 1387.165 | -0.444 | 0.329 | -1.123 | -0.663 | -0.432 | -0.214 | 0.169 |
| Liver → Other interaction | 1.004 | 1.003 | 1.006 | 757.192 | 1155.125 | 0.144 | 0.250 | -0.345 | -0.022 | 0.148 | 0.308 | 0.647 |
| Lymph nodes → Other interaction | 1.007 | 1.005 | 1.016 | 492.449 | 1042.375 | 0.516 | 0.259 | -0.014 | 0.345 | 0.525 | 0.698 | 0.995 |

**Table S22:** Markov Chain Monte Carlo diagnostics for HER2+ disease. Detailed estimation results for parameters relating to the HER2+ group

| Parameter | $\hat{R}$ | PSRF | PSRF<sub>97.5%</sub> | ESSb | ESS<sub>t</sub> | Mean | SD | q2.5 | q25 | q50 | q75 | q97.5 |
| --- | --- | --- | --- | --- | --- | --- | --- | --- | --- | --- | --- | --- |
| Baseline rates |  |  |  |  |  |  |  |  |  |  |  |  |
| log<sub>2</sub> baseline rate Bone | 1.023 | 1.013 | 1.037 | 203.741 | 537.414 | -4.012 | 0.402 | -4.787 | -4.289 | -4.018 | -3.738 | -3.238 |
| log<sub>2</sub> baseline rate Lungs | 1.005 | 1.006 | 1.017 | 594.288 | 1515.587 | -4.245 | 0.464 | -5.167 | -4.558 | -4.231 | -3.922 | -3.371 |
| log<sub>2</sub> baseline rate Liver | 1.010 | 1.005 | 1.014 | 246.832 | 661.986 | -4.841 | 0.549 | -5.945 | -5.211 | -4.816 | -4.457 | -3.819 |
| log<sub>2</sub> baseline rate Lymph nodes | 1.016 | 1.013 | 1.038 | 223.170 | 630.828 | -3.259 | 0.327 | -3.908 | -3.467 | -3.254 | -3.038 | -2.637 |
| log<sub>2</sub> baseline rate Brain | 1.012 | 1.017 | 1.049 | 224.455 | 685.290 | -3.998 | 0.327 | -4.642 | -4.223 | -3.996 | -3.766 | -3.374 |
| log<sub>2</sub> baseline rate Other | 1.020 | 1.019 | 1.057 | 389.825 | 708.816 | -3.989 | 0.417 | -4.828 | -4.273 | -3.983 | -3.707 | -3.173 |
| log<sub>2</sub> first metastasis rate Bone | 1.153 | 1.320 | 2.117 | 27.968 | 21.266 | 3.376 | 2.252 | 0.087 | 2.198 | 3.135 | 4.007 | 10.482 |
| log<sub>2</sub> first metastasis rate Lungs | 1.149 | 1.311 | 2.073 | 27.320 | 21.143 | 4.514 | 2.238 | 1.293 | 3.378 | 4.287 | 5.175 | 11.533 |
| log<sub>2</sub> first metastasis rate Liver | 1.155 | 1.323 | 2.134 | 26.746 | 21.095 | 4.530 | 2.230 | 1.269 | 3.391 | 4.325 | 5.147 | 11.372 |
| log<sub>2</sub> first metastasis rate Lymph nodes | 1.144 | 1.284 | 1.926 | 29.522 | 21.106 | 2.979 | 2.305 | -0.505 | 1.778 | 2.735 | 3.703 | 9.995 |
| log<sub>2</sub> first metastasis rate Brain | 1.153 | 1.313 | 2.082 | 27.177 | 21.156 | 3.824 | 2.227 | 0.592 | 2.670 | 3.615 | 4.434 | 10.831 |
| log<sub>2</sub> first metastasis rate Other | 1.150 | 1.313 | 2.079 | 27.009 | 21.095 | 4.388 | 2.232 | 1.139 | 3.195 | 4.181 | 5.056 | 11.364 |
| Fixed covariate effects |  |  |  |  |  |  |  |  |  |  |  |  |
| Pre-menopausal | 1.022 | 1.022 | 1.063 | 352.891 | 480.598 | 0.564 | 0.222 | 0.139 | 0.418 | 0.560 | 0.703 | 1.015 |
| Unknown menopausal status | 1.009 | 1.011 | 1.033 | 561.017 | 1118.697 | -0.243 | 0.186 | -0.606 | -0.372 | -0.248 | -0.115 | 0.119 |
| Age (pre-menopausal) | 1.007 | 1.007 | 1.017 | 398.469 | 568.335 | 0.129 | 0.667 | -1.192 | -0.311 | 0.118 | 0.581 | 1.450 |
| Age (unknown menopausal status) | 1.013 | 1.008 | 1.022 | 371.480 | 661.639 | 0.199 | 0.677 | -1.153 | -0.255 | 0.195 | 0.662 | 1.539 |
| Age (post-menopausal) | 1.009 | 1.014 | 1.042 | 704.360 | 1256.150 | -0.625 | 0.580 | -1.754 | -1.020 | -0.618 | -0.237 | 0.523 |
| Therapy embedding effects |  |  |  |  |  |  |  |  |  |  |  |  |
| Therapy PC 1 | 1.006 | 1.003 | 1.007 | 1413.576 | 2271.343 | 0.001 | 0.703 | -1.408 | -0.464 | 0.000 | 0.468 | 1.375 |
| Therapy PC 2 | 1.014 | 1.018 | 1.050 | 457.815 | 730.996 | -0.378 | 0.726 | -1.815 | -0.861 | -0.381 | 0.121 | 1.024 |
| Therapy PC 3 | 1.010 | 1.014 | 1.042 | 491.578 | 948.235 | -0.340 | 0.690 | -1.715 | -0.810 | -0.336 | 0.122 | 0.976 |
| Therapy PC 4 | 1.003 | 1.001 | 1.005 | 994.644 | 1737.850 | 0.119 | 0.694 | -1.256 | -0.340 | 0.114 | 0.574 | 1.501 |
| Therapy PC 5 | 1.003 | 1.001 | 1.004 | 1274.448 | 2018.531 | -0.047 | 0.717 | -1.454 | -0.538 | -0.051 | 0.445 | 1.389 |
| Therapy PC 6 | 1.014 | 1.017 | 1.029 | 384.949 | 632.485 | -0.245 | 0.698 | -1.627 | -0.713 | -0.238 | 0.221 | 1.093 |
| Therapy PC 7 | 1.002 | 1.000 | 1.000 | 1490.212 | 2375.119 | 0.353 | 0.700 | -1.021 | -0.119 | 0.340 | 0.832 | 1.731 |
| Therapy PC 8 | 1.002 | 1.002 | 1.007 | 685.124 | 1226.465 | 0.192 | 0.700 | -1.182 | -0.280 | 0.198 | 0.654 | 1.558 |
| Therapy PC 9 | 1.012 | 1.007 | 1.020 | 563.015 | 1188.549 | 0.208 | 0.718 | -1.216 | -0.287 | 0.215 | 0.712 | 1.560 |
| Therapy PC 10 | 1.005 | 1.005 | 1.014 | 909.705 | 1791.050 | -0.275 | 0.713 | -1.629 | -0.765 | -0.282 | 0.213 | 1.156 |
| Interaction effects |  |  |  |  |  |  |  |  |  |  |  |  |
| Lungs → Bone interaction | 1.008 | 1.010 | 1.028 | 339.784 | 1259.178 | 0.881 | 0.373 | 0.132 | 0.630 | 0.891 | 1.144 | 1.579 |
| Liver → Bone interaction | 1.018 | 1.006 | 1.018 | 337.239 | 1216.359 | 0.714 | 0.363 | -0.027 | 0.475 | 0.723 | 0.960 | 1.402 |
| Lymph nodes → Bone interaction | 1.006 | 1.004 | 1.009 | 683.162 | 1351.536 | 0.055 | 0.394 | -0.757 | -0.205 | 0.068 | 0.319 | 0.826 |
| Other → Bone interaction | 1.028 | 1.006 | 1.019 | 274.366 | 596.103 | 0.440 | 0.384 | -0.338 | 0.183 | 0.445 | 0.696 | 1.183 |
| Bone → Lungs interaction | 1.008 | 1.003 | 1.010 | 645.523 | 1316.970 | 0.071 | 0.431 | -0.782 | -0.227 | 0.070 | 0.365 | 0.922 |
| Liver → Lungs interaction | 1.014 | 1.012 | 1.033 | 476.036 | 1290.523 | 0.150 | 0.425 | -0.682 | -0.138 | 0.150 | 0.441 | 0.980 |
| Lymph nodes → Lungs interaction | 1.003 | 1.005 | 1.013 | 631.871 | 1128.399 | -0.073 | 0.444 | -0.954 | -0.373 | -0.075 | 0.221 | 0.764 |
| Other → Lungs interaction | 1.007 | 1.008 | 1.024 | 630.439 | 1131.967 | 1.184 | 0.434 | 0.341 | 0.889 | 1.178 | 1.488 | 2.017 |
| Bone → Liver interaction | 1.015 | 1.013 | 1.037 | 421.798 | 813.182 | 0.229 | 0.487 | -0.713 | -0.090 | 0.223 | 0.546 | 1.210 |
| Lungs → Liver interaction | 1.007 | 1.002 | 1.006 | 417.619 | 938.177 | 1.181 | 0.455 | 0.288 | 0.877 | 1.179 | 1.492 | 2.081 |
| Lymph nodes → Liver interaction | 1.003 | 1.002 | 1.004 | 748.686 | 1172.606 | 0.295 | 0.437 | -0.586 | -0.001 | 0.299 | 0.598 | 1.140 |
| Other → Liver interaction | 1.010 | 1.015 | 1.039 | 499.756 | 604.079 | 0.623 | 0.453 | -0.305 | 0.329 | 0.628 | 0.928 | 1.496 |
| Bone → Lymph nodes interaction | 1.003 | 1.002 | 1.004 | 808.328 | 1221.074 | -0.842 | 0.408 | -1.662 | -1.120 | -0.828 | -0.557 | -0.064 |
| Lungs → Lymph nodes interaction | 1.007 | 1.007 | 1.021 | 871.945 | 1442.864 | 0.391 | 0.316 | -0.247 | 0.182 | 0.388 | 0.614 | 0.983 |
| Liver → Lymph nodes interaction | 1.007 | 1.008 | 1.025 | 662.080 | 1008.513 | 0.257 | 0.333 | -0.399 | 0.033 | 0.257 | 0.489 | 0.889 |
| Other → Lymph nodes interaction | 1.006 | 1.009 | 1.027 | 369.444 | 772.227 | 0.522 | 0.333 | -0.156 | 0.299 | 0.531 | 0.745 | 1.163 |
| Bone → Brain interaction | 1.004 | 1.003 | 1.010 | 595.696 | 1017.689 | 0.336 | 0.274 | -0.212 | 0.152 | 0.337 | 0.523 | 0.871 |
| Lungs → Brain interaction | 1.002 | 1.002 | 1.007 | 929.146 | 1410.181 | 0.886 | 0.277 | 0.332 | 0.703 | 0.893 | 1.072 | 1.414 |
| Liver → Brain interaction | 1.010 | 1.013 | 1.039 | 478.387 | 992.435 | 0.519 | 0.273 | -0.023 | 0.329 | 0.522 | 0.713 | 1.045 |
| Lymph nodes → Brain interaction | 1.008 | 1.008 | 1.023 | 665.298 | 1055.612 | 0.010 | 0.281 | -0.524 | -0.181 | 0.006 | 0.201 | 0.570 |
| Other → Brain interaction | 1.010 | 1.012 | 1.035 | 628.537 | 1018.556 | 0.290 | 0.309 | -0.339 | 0.085 | 0.297 | 0.507 | 0.871 |
| Bone → Other interaction | 1.004 | 1.002 | 1.005 | 818.723 | 1403.671 | -0.031 | 0.383 | -0.796 | -0.282 | -0.028 | 0.223 | 0.713 |
| Lungs → Other interaction | 1.003 | 1.004 | 1.013 | 990.773 | 1254.123 | 0.259 | 0.392 | -0.525 | 0.003 | 0.260 | 0.522 | 1.027 |
| Liver → Other interaction | 1.021 | 1.027 | 1.079 | 382.918 | 549.106 | -0.180 | 0.403 | -0.992 | -0.447 | -0.166 | 0.088 | 0.594 |
| Lymph nodes → Other interaction | 1.009 | 1.008 | 1.020 | 486.648 | 807.035 | 0.352 | 0.374 | -0.377 | 0.118 | 0.348 | 0.599 | 1.093 |

**Table S23:**
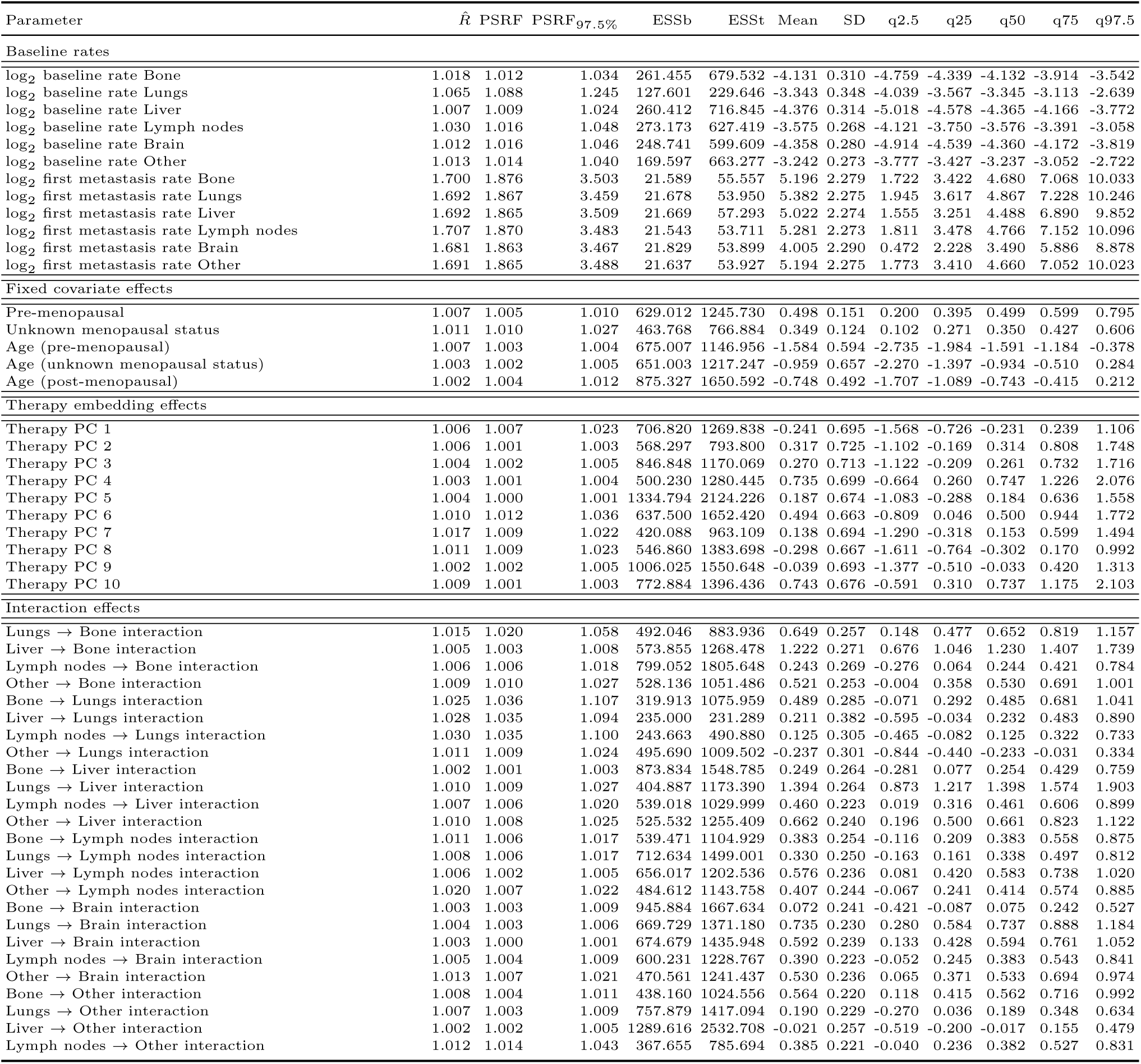
Markov Chain Monte Carlo diagnostics for triple-negative disease. Detailed estimation results for parameters relating to the TNBC group

